# Hidden in the Night: Wearable Sleep Assessment of Nocturnal Hypoglycaemia in Type 1 Diabetes

**DOI:** 10.64898/2026.01.22.26344161

**Authors:** Ashwaq Alsuhaymi, Paul Nutter, Hood Thabit, Simon Harper

## Abstract

**Background:** Nocturnal hypoglycaemia (NH) is a common and challenging complication in Type 1 Diabetes (T1D), disrupting blood glucose control and sleep physiology. Its real-world impact on sleep architecture remains poorly characterised. Consumer wearables offer a way to examine these associations under free-living conditions, providing detailed insight into behavioural and physiological responses to nocturnal blood glucose fluctuations. This study aims to assess how wearable-derived sleep metrics and physiological features could be used as indicators of NH, including the effects of how low blood glucose levels fall during hypoglycaemic events and the associated pre-event changes.

**Methods:** We conducted a comparative observational analysis of paired continuous glucose monitoring (CGM) and Garmin smartwatch data collected over 12 weeks from 17 adults with T1D. Nights were categorised as normoglycaemia, hyperglycaemia, or hypoglycaemia Level 1 (*≥*3.1 and *<*3.9 mmol/L), and hypoglycaemia Level 2 (*<*3.0 mmol/L). Thirteen sleep metrics, including total sleep time, wake after sleep onset (WASO), sleep-stage proportions, fragmentation indices, and physiological features such as heart rate, were compared using non-parametric tests. Pre-hypoglycaemic event analyses examined 60-minute and 15-minute windows preceding hypoglycaemia to identify early deviations in sleep and physiological metrics.

**Results:** Across 573 nights, 17.5% involved Level 1 and 7.3% Level 2 hypoglycaemia. Level 2 hypoglycaemia was associated with 31 minutes less wakefulness, 17–25 minutes more REM, and up to 74% more deep sleep compared with normo-glycaemic nights. Sleep efficiency increased during hypoglycaemic events despite greater fragmentation. Pre-hypoglycaemic episode analyses revealed shorter awake and light-sleep bouts, as well as a 9.8% higher heart rate, preceding Level 2 episodes.

**Conclusions:** Wearable-derived sleep and physiological signals reveal clear intraindividual changes both before and during NH. Our findings indicate that Level 2 episodes are associated with deeper sleep and reduced behavioural arousal, suggesting that CGM alarms may be less effective at waking individuals during level2 NH. By characterising pre-hypoglycaemic changes that differ based on hypoglycaemia level, this work provides preliminary evidence for personalised, wearable-based early-warning systems. Such approaches could help distinguish nocturnal hypoglycaemic events and support more effective alerting, particularly in settings with limited or no access to CGM.

**Author Summary:** *Why was this study done?:* People with Type 1 Diabetes (T1D) frequently experience nocturnal hypoglycaemia (low blood glucose at night), a dangerous event that often goes unnoticed because individuals are less able to recognise symptoms or wake up during sleep. These events also disrupt sleep in ways that are not well characterised under real-world conditions. Limited access to continuous glucose monitoring (CGM), especially in low- and middle-income countries, highlights the need for affordable alternatives to ensure nighttime safety.

*What did we do and find?:* Using more than 500 nights of paired smartwatch and CGM data, we investigated how sleep features change when blood glucose levels fall overnight. We found that hypoglycaemic nights show distinct alterations in sleep architecture, including increased REM and deep sleep, and greater micro-fragmentation. A key finding was that Level 2 hypoglycaemia was associated with deeper sleep and reduced wakefulness. This pattern indicates that individuals may be less likely to awaken during more severe events, even when alarms are present. Pre-hypoglycaemic episode analysis revealed additional early-warning signals, such as shorter awake and light-sleep bouts and elevated heart rate, before level 2 hypoglycaemia occurred.

*What do these findings mean?:* Smartwatches can capture sleep-based changes that appear before and during nocturnal hypoglycaemia. Because deeper sleep during Level 2 episodes may reduce responsiveness to CGM alerts, these results suggest that current alarm approaches could be improved by incorporating sleep features alongside glucose data. Such sleep-informed detection may enhance the reliability of hypoglycaemia alerts, reduce missed events during deep sleep, and provide a foundation for low-cost early-warning systems in settings where CGM is unavailable or unaffordable. Further research is needed in larger and more diverse populations, but this work provides early evidence that wearable-derived sleep features can meaningfully strengthen nocturnal hypoglycaemia detection.

## Introduction

Diabetes management is essential for individuals with T1D, yet unpredictable glucose fluctuations continue to increase both acute and long-term complications [1]. Among these fluctuations, hypoglycaemia remains one of the most persistent barriers to effective self-management. NH is particularly concerning, occurring during sleep when awareness and behavioural defences are diminished and often going undetected until morning. Globally, an estimated 8.7 million people live with T1D in 2021, with adults aged 20 years and older accounting for 64% of all cases [2]. NH affects up to 50% of adults with T1D during overnight monitoring, with 25-90% of episodes remaining asymptomatic, and these events account for nearly half of all severe hypoglycaemic episodes occurring during sleep [3]. Large observational studies indicate that the dead-in-bed syndrome accounts for approximately 5-6% of deaths in individuals with T1D under 40 years of age, in which nocturnal hypoglycaemia is identified as the principal precipitating factor [4, 5]. These events extend beyond transient glucose dips: recurrent NH disrupts sleep architecture, reducing restorative deep sleep and rapid eye movement (REM) sleep while increasing nocturnal wakefulness [6–10]. Such alterations impair growth hormone release, cognitive performance, and next-day metabolic stability, creating a self-reinforcing cycle of dysglycaemia and reduced quality of life [11–15]. Understanding how nocturnal glucose patterns interact with sleep physiology is essential for improving both metabolic and mental health outcomes in adults with T1D.

CGM has substantially improved diabetes care by providing real-time interstitial blood glucose data, trend alerts, and insights into daily variability [16–19]. Yet, CGM use and adoption remain inconsistent across patient populations. Many adults discontinue CGM or disable alerts due to device cost, alarm fatigue, sensor inaccuracies resulting in false alerts, and perceived loss of autonomy [18–20]. These barriers are further amplified in low- and middle-income countries (LMICs), where access to CGM is constrained by high costs, limited clinical training, and inadequate insurance coverage [21–23]. In many LMIC settings, individuals rely primarily on intermittent capillary blood glucose checks, often only a few times per week, leaving NH largely undetected [21]. Conversely, adverse behaviours to avoid NH (such as pre-bed or night-time snacking) may also contribute to elevated glycated haemoglobin A1c (HbA1c) levels and increased risk of chronic complications [24–26]. These limitations highlight the need for accessible, low-cost physiological monitoring tools.

Recent advances in consumer-grade wearable technology offer a promising avenue for addressing these challenges [26]. Affordable smartwatches and fitness trackers can continuously capture physiological signals that reflect glycaemic fluctuations, including heart rate (HR), heart-rate variability (HRV), skin temperature, respiratory rate, movement and sleep [27–30]. Machine-learning models trained on such multimodal data have shown encouraging accuracy for detecting hypoglycaemic events [30–34]. Cardiac and movement-based features remain informative during sleep, suggesting that nocturnal detection may be a particular strength of wearable-based systems [35–37]. Beyond static cohort-based prediction models, adaptive systems represent an important evolution, enabling models to refine personalised thresholds based on each individual’s physiological and behavioural patterns [38, 39]. Through repeated nightly observations, these systems may identify subtle, individual-specific markers that precede hypoglycaemia. This approach opens the possibility of low-cost, non-invasive, and scalable nocturnal detection using devices already owned by millions worldwide [40], with particular relevance to LMICs where CGM remains inaccessible to most individuals with T1D.

While automated detection of hypoglycaemia from smartwatch data is advancing [28], the interaction between sleep–wake behaviour and nocturnal blood glucose patterns remains relatively understudied. Accurate identification of sleep–wake states is essential for both physiological interpretation and algorithm refinement [41]. Evidence shows that traditional fixed clock-time windows (e.g., 00:00–06:00) underestimate NH compared with personalised, wearable-defined sleep intervals [42]. Although modern multi-sensor wearables achieve moderate accuracy for sleep staging relative to polysomnography [36, 43, 44], their continuous multi-sensor analysis enables clinically meaningful interpretation of nocturnal physiology in free-living conditions. Integrating adaptive modelling into sleep analysis may further personalise detection by learning each individual’s typical sleep structure and identifying deviations that could be associated with elevated hypoglycaemic risk.

This study addresses an important and unmet clinical need. While prior research has explored wearable-based detection of hypoglycaemia, no studies have examined how wearable-derived sleep architecture and physiology change before and during NH in free-living adults with T1D, or how these pre-hypoglycaemic episode changes differ by hypoglycaemic level. To address this, our work integrates: (1) synchronised CGM and smartwatch data, (2) detailed wearable-derived sleep architecture, (3) pre-hypoglycaemic episode physiological and sleep-stage dynamics, and (4) severity-based analysis of hypoglycaemia (level 1 vs. level 2). This combination provides the first systematic characterisation of NH within the context of real-world sleep, offering foundational insight for personalised and adaptive early-warning systems.

The primary objective of this study is to characterise the association between nocturnal hypoglycaemic episodes and sleep behaviour derived from smartwatch data, examining both within-individual night-to-night variability and between-individual differences. The secondary objectives are to quantify sleep features that change before NH occurs and determine how these pre-hypoglycaemic episode alterations differ by NH episode level, as well as identify key physiological markers associated with these events. Together, these objectives address a critical gap in digital diabetes care and demonstrate how consumer-grade wearables may support personalised blood glucose monitoring and prediction, particularly in settings where traditional CGM systems remain inaccessible.

## Results

A total of 663 nights with ISG data were initially available for analysis. After aligning and merging these with corresponding sleep-stage data, 573 nights contained complete and synchronised records. Of these, 174 nights (30.4%) were classified as normoglycaemia, 257 nights (44.9%) as hyperglycaemia, 100 nights (17.5%) as hypoglycaemia Level 1, and 42 nights (7.3%) as hypoglycaemia Level 2. Across these categories, blood glucose variability was up to three times higher for hypoglycaemic nights compared with normoglycaemia nights. The highest variability was observed in Hypoglycaemia Level 2 nights (coefficient of variation = 32.1%).

Sleep quality and efficiency metrics also varied according to glycaemic classification. Nights with hyperglycaemia exhibited the lowest overall sleep quality (mean score = 60.4) and greater disruption, characterised by 27.6% more WASO and a significantly higher number of awakenings (p *<*0.001). In contrast, hypoglycaemic nights showed higher sleep efficiency (91–92% compared to 88% in normoglycaemic nights) and longer total sleep duration, with the most pronounced increases observed during hyperglycaemic and hypoglycaemia Level 2 nights. Hypoglycaemia Level 2 nights exhibited more than 50% increase in deep sleep (74% recorded increase), while Hypoglycaemia Level 1 showed a 24% increase in deep sleep and a 2.5-fold higher proportion of REM sleep compared to normoglycaemic nights. The most pronounced increase in deep sleep was observed during Severe Hypoglycaemia (Level 2), with a mean duration of 222.5 minutes (approximately three hours). Figure 1 summarises, for each participant, the percentage distribution of ISG ranges and sleep stages, with bars normalised to 100% per subject.

**Fig 1.**
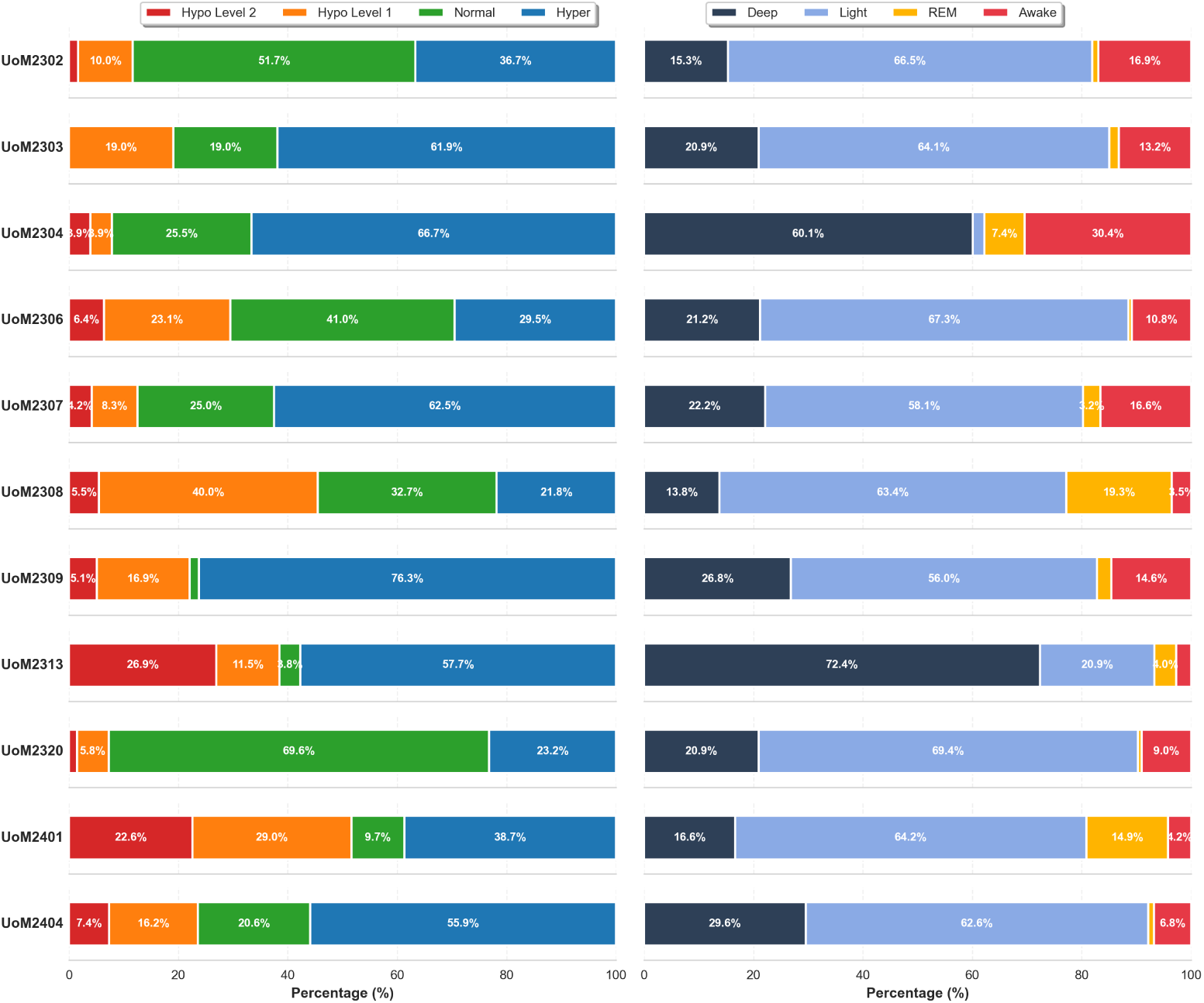
Participant-level distributions shown as stacked horizontal bar charts. Left: glucose time-in-range (Hypo Level 2, Hypo Level 1, Normal, Hyper). Right: sleep-stage composition (Deep, Light, REM, Awake). Each row corresponds to one participant (UoM IDs), and bars sum to 100% along the x-axis (Percentage).

### Sleep Duration and Regularity

Sleep duration and regularity patterns were analysed across the 11 participants, considering nocturnal hypoglycaemic episodes. The mean sleep duration was 8.67 *±* 1.05 hours, with all participants achieving adequate sleep duration (*≥* 7 hours) and 82% (*n* = 9) falling within the recommended 7–9 hour range. Two participants (18%) exhibited extended sleep durations exceeding 9 hours (range: 9.23–11.54 hours). The inter-individual variability in sleep timing and duration across the study cohort is illustrated in Figure 2. Sleep regularity, assessed via the circular standard deviation of sleep onset times, demonstrated considerable inter-individual variability (mean = 1.15 *±* 0.48 hours). Forty-five percent of participants (*n* = 5) maintained highly regular sleep schedules with onset variability *<* 1 hour, while another 45% (*n* = 5) showed moderate regularity (1–2 hours of variability). One participant (UoM2313, 9%) exhibited irregular sleep patterns with *>* 2 hours of variability.

**Fig 2.**
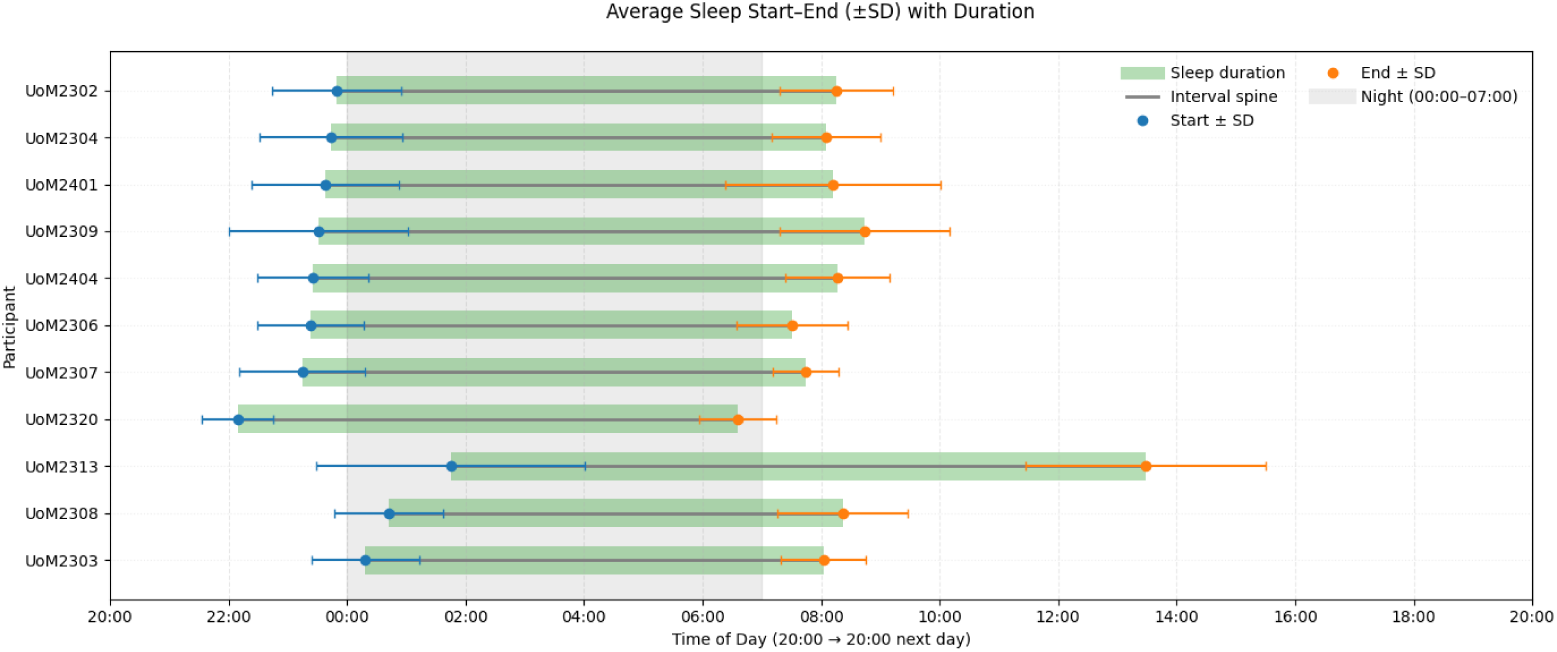
Average sleep start and end times (mean *±* SD) with duration for all participants. Each horizontal bar represents the mean sleep interval per participant, with shaded regions indicating standard deviation around the mean sleep duration. Blue and orange markers denote the mean start and end times, respectively, while the grey area represents the typical night period (00:00–07:00).

Hypoglycaemic episodes occurred on 24.8% of all monitored nights, with participants experiencing episodes on 20.0% *±* 16.7% of their individual nights (range: 0.0%–46.3%). Participants with the highest hypoglycaemic burden (UoM2401: 45.0%; UoM2308: 46.3%) demonstrated adequate sleep duration but moderate schedule variability, whereas the participant with regular sleep (UoM2320: *±* 0.60 hours) experienced the lowest hypoglycaemic rate (5.8%).

### Sleep Features Characterising Hypoglycaemic Nights

Kruskal–Wallis tests revealed significant differences across glycaemic categories for 31.1% of sleep features examined. The most pronounced differences were observed in fundamental sleep architecture metrics, including sleep quality score (*H* = 60.35, *p* = 4.95 *×* 10*^−^*^13^), time spent awake (*H* = 54.85, *p* = 7.40 *×* 10*^−^*^12^), wake after sleep onset (WASO; *H* = 54.83, *p* = 7.47 *×* 10*^−^*^12^), and deep sleep duration (*H* = 42.49, *p* = 3.16 *×* 10*^−^*^9^).

#### Reduced Wakefulness

Pairwise comparisons revealed that hypoglycaemic nights were characterised by significantly less time spent awake compared to both hyperglycaemic and normoglycaemic nights (Supplementary materials: Table SD). Hypoglycaemic Level 2 nights showed a median of 42.0 minutes awake, compared to 73.0 minutes during hyperglycaemic nights (median difference = *−*31.0 min, *p* = 1.73 *×* 10*^−^*^9^, Cliff’s *δ* = *−*0.360, medium effect) and 64.0 minutes during normoglycaemic nights (median difference = *−*22.0 min, *p* = 2.42 *×* 10*^−^*^4^, Cliff’s *δ* = *−*0.267, small effect).

Hypoglycaemia Level 1 nights similarly demonstrated reduced wakefulness, with a median of 49.0 minutes awake, significantly lower than hyperglycaemic nights (median difference = *−*24.0 min, *p* = 1.10 *×* 10*^−^*^8^, Cliff’s *δ* = *−*0.263, small effect) and normoglycaemic nights (median difference = *−*15.0 min, *p* = 9.89 *×* 10*^−^*^3^, Cliff’s *δ* = *−*0.160, small effect). WASO showed an identical pattern of reduction during hypoglycaemic nights, with the same statistical parameters, as presented in Figure 3.

**Fig 3.**
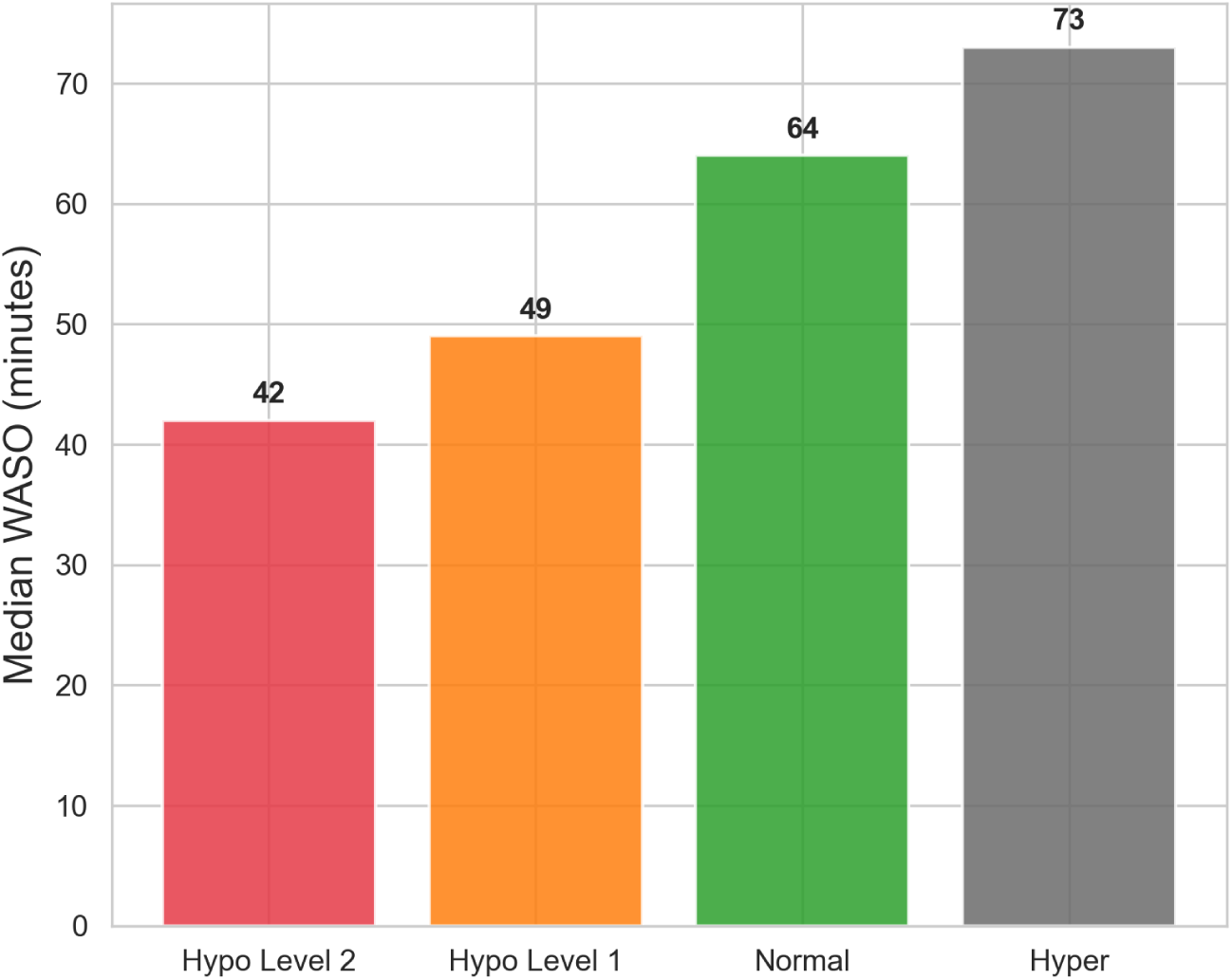
Median wake after sleep onset (WASO) across glycaemic categories in minutes.

#### REM Sleep Alterations

Pairwise comparisons revealed that hypoglycaemic nights were associated with significantly greater REM sleep duration compared to normoglycaemic nights. Hypo Level 2 nights had a median REM duration of 21.0 minutes, compared to 4.0 minutes in normoglycaemic nights (median difference = 17.0 min, *p* = 1.79 *×* 10*^−^*^6^, Cliff’s *δ* = 0.347, medium effect) and 9.0 minutes in hyperglycaemic nights (median difference = 12.0 min, *p* = 9.12 *×* 10*^−^*^3^, Cliff’s *δ* = 0.156, small effect) (Fig. 4).

**Fig 4.**
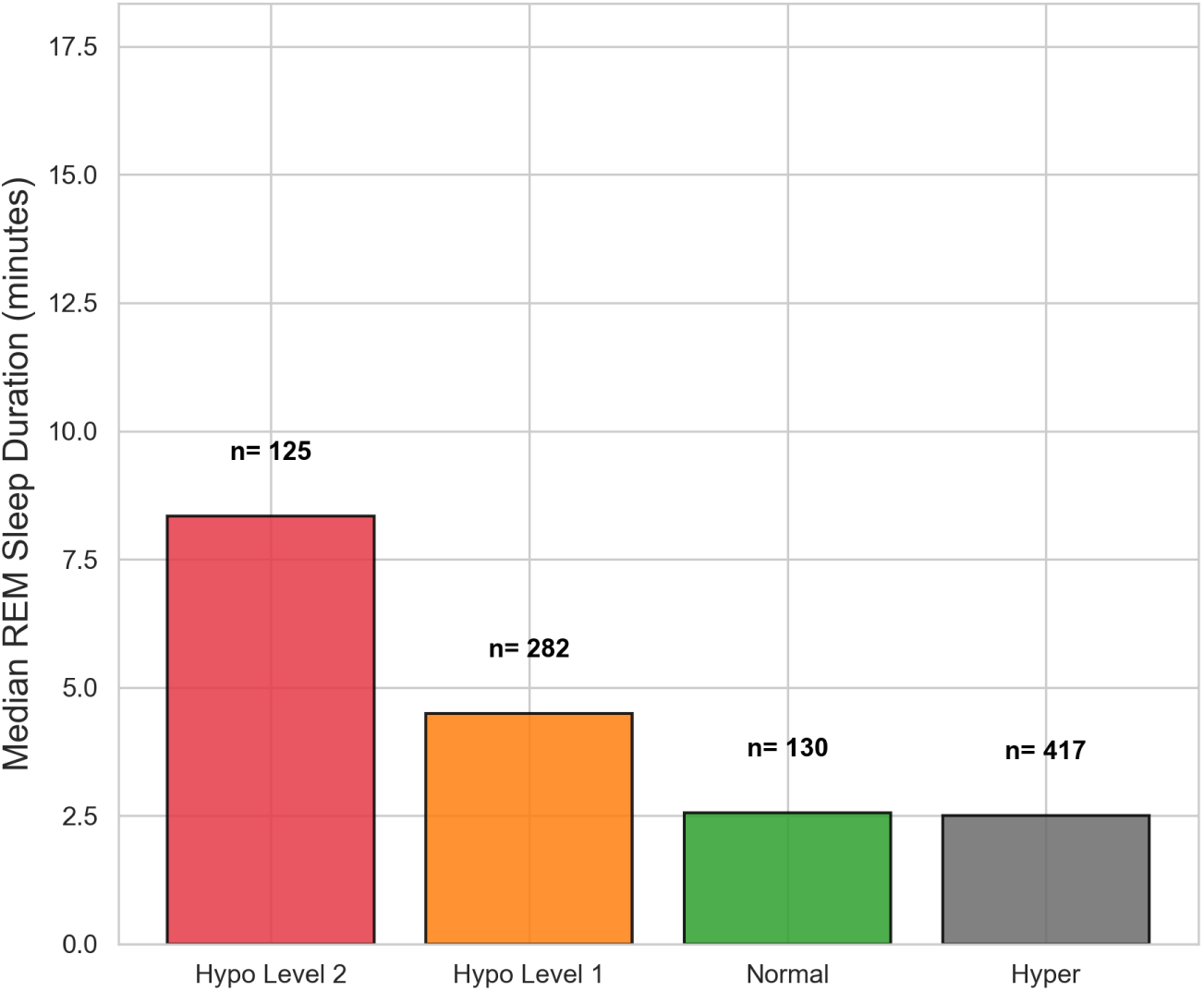
Median REM sleep duration across glycaemic night types. Bar plot showing median REM sleep duration (minutes) across hypoglycaemic (Level 2, Level 1), normoglycaemic, and hyperglycaemic nights.

Hypo Level 1 nights similarly showed elevated REM sleep, with a median of 11.0 minutes, significantly higher than normoglycaemic nights (median difference = 7.0 min, *p* = 1.36 *×* 10*^−^*^6^, Cliff’s *δ* = 0.299, small effect).

#### Sleep Fragmentation and Stability Indices

Kruskal-Wallis tests, presented in Table 1, revealed significant differences in the awake fragmentation index across glycaemic classifications (*H* = 14.84, *p* = 1.96 *×* 10*^−^*^3^), indicating that the pattern and frequency of awakenings varied according to nocturnal blood glucose level. Pairwise comparisons showed that hypoglycaemic nights exhibited significantly higher awake fragmentation than both hyperglycaemic and normoglycaemic nights. Hypo Level 2 nights had a median awake fragmentation index of 7.500, compared to 1.352 during hyperglycaemic nights (median difference = 6.148, *p* = 2.22 *×* 10*^−^*^3^,

**Table 1.**
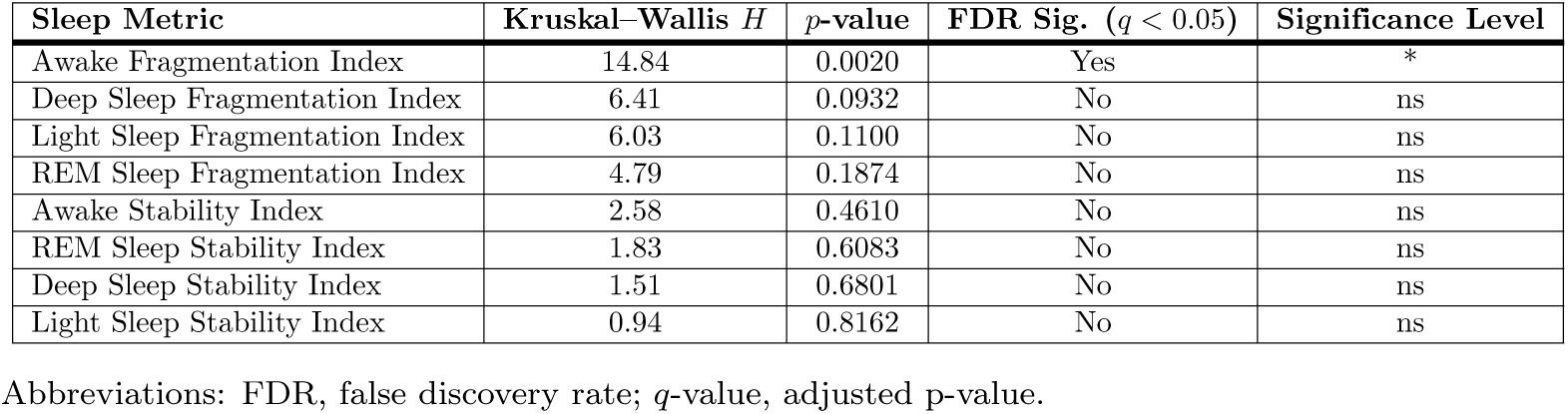
Kruskal–Wallis tests for fragmentation and stability indices across glycaemic night categories.

Cliff’s *δ* = 0.536, large effect) and 1.096 during normoglycaemic nights (median difference = 6.404, *p* = 1.43 *×* 10*^−^*^3^, Cliff’s *δ* = 0.599, large effect).

Hypoglycaemia Level 1 nights similarly showed elevated awake fragmentation, with a median of 7.008, significantly higher than hyperglycaemic nights (median difference = 5.656, *p* = 1.45 *×* 10*^−^*^2^, Cliff’s *δ* = 0.285, small effect) and normoglycaemic nights (median difference = 5.912, *p* = 2.87 *×* 10*^−^*^2^, Cliff’s *δ* = 0.292, small effect).

Fragmentation indices for deep, light, and REM sleep stages did not differ significantly across glycaemic states (Deep: *H* = 6.41, *p* = 0.093; Light: *H* = 6.03, *p* = 0.110; REM: *H* = 4.79, *p* = 0.187), indicating that fragmentation during hypoglycaemic nights was specific to the awake stage rather than affecting all sleep stages uniformly. Similarly, sleep-stage stability indices (Awake, Deep, Light, REM) showed no significant differences across glycaemic categories (all *p >* 0.05), suggesting that whilst the amount and fragmentation of sleep stages varied during hypoglycaemic nights, the inherent stability of individual stages remained unchanged.

Stage transition metrics showed increased frequency during hypoglycaemic nights. The overall Kruskal–Wallis tests identified significant group effects across multiple stage–transition metrics. Hypoglycaemic nights showed consistent disruptions in sleep–stage dynamics compared with both normoglycaemic and hyperglycaemic nights. Pairwise comparisons revealed that hypoglycaemic nights, particularly Level 2 hypoglycaemia, exhibited the most pronounced alterations. Both hypoglycaemia Level 1 and Level 2 nights showed significantly more awake to deep transitions compared with normoglycaemic nights (U = 12 821.5, *p* = 0.00062, FDR-corrected *p* = 0.0037, and U = 21 717.5, *p* = 0.0116, respectively), as shown in table 2.

**Table 2.**
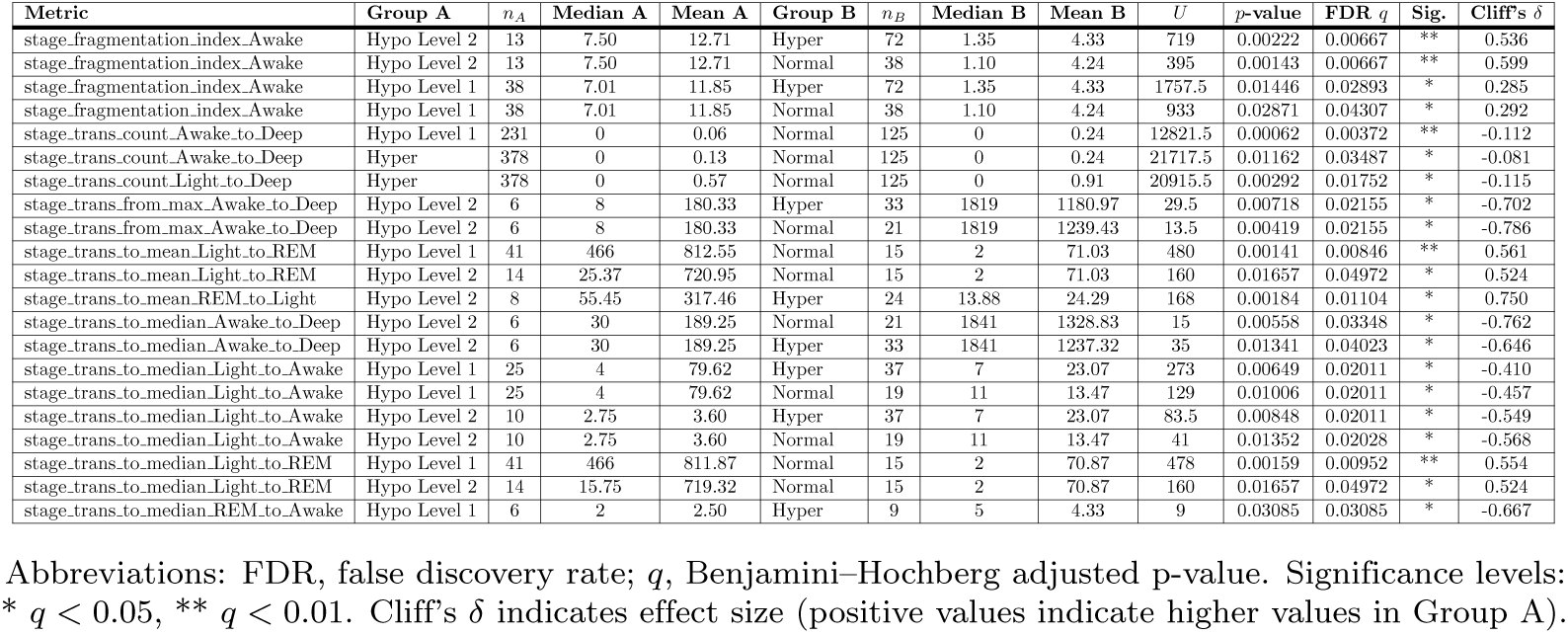
Pairwise Mann-Whitney U tests for sleep fragmentation and transition metrics across glycaemic night groups.

#### Physiological Parameters

Pairwise Mann–Whitney U tests revealed significant differences in multiple physiological metrics across glycaemic groups (Table 3). Nocturnal step count data revealed significantly higher physical activity during hypoglycaemic nights compared to normoglycaemic and hyperglycaemic nights. Step counts were elevated during hypoglycaemic episodes (Hypo vs. Norm: *U* = 156,789, *p <* 0.001; Hypo vs. Hyper: *U* = 112,345, *p <* 0.001), with within-participant comparisons showing a stronger effect. When step data occurring immediately after hypoglycaemia onset were excluded, nocturnal activity levels were substantially reduced, demonstrating that increased movement was concentrated around hypoglycaemic episodes.

**Table 3.** Pairwise Mann-Whitney U tests for physiological heart rate, step count, and stress metrics across glycaemic groups.

| Metric | Group A | $n_A$ | Median A | Mean A | Group B | $n_B$ | Median B | Mean B | $U$ | $p$-value | FDR $q$ | Sig. | Cliff's $\delta$ |
| --- | --- | --- | --- | --- | --- | --- | --- | --- | --- | --- | --- | --- | --- |
| physio_heart_rate_mean | Hyper | 257 | 84.99 | 85.48 | Hypo Level 1 | 100 | 80.73 | 80.68 | 16200.0 | 1.31e-04 | 7.84e-04 | *** | 0.261 |
| physio_heart_rate_median | Hyper | 257 | 84.00 | 84.53 | Hypo Level 1 | 100 | 79.50 | 79.47 | 16094.5 | 2.11e-04 | 1.27e-03 | ** | 0.252 |
| physio_heart_rate_n | Normal | 174 | 31 | 30.01 | Hyper | 257 | 34 | 37.86 | 14118.0 | 7.78e-11 | 4.67e-10 | *** | -0.369 |
| physio_heart_rate_n | Normal | 174 | 31 | 30.01 | Hypo Level 1 | 100 | 34 | 37.09 | 5792.5 | 3.96e-06 | 1.19e-05 | *** | -0.334 |
| physio_heart_rate_n | Normal | 174 | 31 | 30.01 | Hypo Level 2 | 42 | 34.5 | 38.40 | 2416.5 | 6.47e-04 | 1.29e-03 | ** | -0.339 |
| physio_step_count_n | Normal | 174 | 31 | 30.01 | Hyper | 257 | 34 | 37.86 | 14118.0 | 7.78e-11 | 4.67e-10 | *** | -0.369 |
| physio_step_count_n | Normal | 174 | 31 | 30.01 | Hypo Level 1 | 100 | 34 | 37.09 | 5792.5 | 3.96e-06 | 1.19e-05 | *** | -0.334 |
| physio_step_count_n | Normal | 174 | 31 | 30.01 | Hypo Level 2 | 42 | 34.5 | 38.40 | 2416.5 | 6.47e-04 | 1.29e-03 | ** | -0.339 |
| physio_stress_level_value_max | Normal | 174 | 22.00 | 28.22 | Hyper | 257 | 36.00 | 41.18 | 12579.0 | 1.27e-14 | 7.65e-14 | *** | -0.437 |
| physio_stress_level_value_max | Normal | 174 | 22.00 | 28.22 | Hypo Level 1 | 100 | 34.33 | 39.59 | 5170.5 | 2.28e-08 | 6.83e-08 | *** | -0.406 |
| physio_stress_level_value_max | Normal | 174 | 22.00 | 28.22 | Hypo Level 2 | 42 | 44.83 | 44.83 | 1653.0 | 3.71e-08 | 7.42e-08 | *** | -0.548 |
| physio_stress_level_value_mean | Normal | 174 | 10.05 | 11.77 | Hyper | 257 | 12.69 | 15.14 | 14745.0 | 1.97e-09 | 1.18e-08 | *** | -0.341 |
| physio_stress_level_value_mean | Normal | 174 | 10.05 | 11.77 | Hypo Level 2 | 42 | 14.10 | 15.66 | 1965.0 | 3.40e-06 | 1.02e-05 | *** | -0.462 |
| physio_stress_level_value_mean | Normal | 174 | 10.05 | 11.77 | Hypo Level 1 | 100 | 11.71 | 13.75 | 6379.0 | 2.38e-04 | 4.76e-04 | *** | -0.267 |
| physio_stress_level_value_median | Normal | 174 | 10.00 | 11.05 | Hyper | 257 | 11.67 | 13.50 | 17069.0 | 3.05e-05 | 1.83e-04 | *** | -0.237 |
| physio_stress_level_value_median | Normal | 174 | 10.00 | 11.05 | Hypo Level 2 | 42 | 12.17 | 13.78 | 2403.5 | 5.84e-04 | 1.75e-03 | ** | -0.342 |
| physio_stress_level_value_median | Normal | 174 | 10.00 | 11.05 | Hypo Level 1 | 100 | 11.33 | 11.92 | 7004.5 | 7.26e-03 | 1.45e-02 | * | -0.195 |
| physio_stress_level_value_n | Normal | 174 | 31 | 30.01 | Hyper | 257 | 34 | 37.86 | 14118.0 | 7.78e-11 | 4.67e-10 | *** | -0.369 |
| physio_stress_level_value_n | Normal | 174 | 31 | 30.01 | Hypo Level 1 | 100 | 34 | 37.09 | 5792.5 | 3.96e-06 | 1.19e-05 | *** | -0.334 |
| physio_stress_level_value_n | Normal | 174 | 31 | 30.01 | Hypo Level 2 | 42 | 34.5 | 38.40 | 2416.5 | 6.47e-04 | 1.29e-03 | ** | -0.339 |
| physio_stress_level_value_sd | Normal | 174 | 5.52 | 7.09 | Hyper | 257 | 8.09 | 10.39 | 12995.0 | 1.58e-13 | 9.50e-13 | *** | -0.419 |
| physio_stress_level_value_sd | Normal | 174 | 5.52 | 7.09 | Hypo Level 1 | 100 | 7.50 | 9.78 | 5391.0 | 1.61e-07 | 4.83e-07 | *** | -0.380 |
| physio_stress_level_value_sd | Normal | 174 | 5.52 | 7.09 | Hypo Level 2 | 42 | 9.05 | 10.76 | 1969.0 | 3.59e-06 | 7.18e-06 | *** | -0.461 |

Abbreviations: FDR, false discovery rate; $q$ , Benjamini-Hochberg adjusted p-value. Significance levels: \* $q < 0.05$ , \*\* $q < 0.01$ , \*\*\* $q < 0.001$ . Cliff's $\delta$ represents effect size (negative values indicate higher values in Group B).

Heart rate patterns differed across glycaemic states. Mean and median heart rates were lower during hypoglycaemic nights compared to hyperglycaemic nights (mean difference = 4.8 bpm, *p <* 0.001, Cliff’s *δ* = 0.26, small effect). However, the number of heart rate events was higher during both hyperglycaemic and hypoglycaemic nights relative to normoglycaemic nights (*p <* 0.001, *δ ≈* 0.33–0.37, medium effect).

Stress-related measures showed a clear pattern across glycaemic categories. Maximum stress values increased progressively from normoglycaemic to hyperglycaemic to Hypoglycaemic Level 2 nights (median = 22.0, 36.0, and 44.8, respectively; *p <* 10*^−^*^8^, Cliff’s *δ* = 0.55, large effect). Mean and median stress levels followed the same pattern (*p <* 0.001) with small-to-medium effect sizes (*δ* = 0.27–0.46). Stress variability (SD) was significantly greater during both hyperglycaemic and hypoglycaemic nights compared to normoglycaemic nights (*p <* 10*^−^*^6^, *δ ≈* 0.4, medium effect).

### Pre-Hypoglycaemic Episode Analysis

To further characterise the temporal relationship between physiological changes and hypoglycaemic episodes, we examined 750 pre-hypoglycaemic episode windows (60 minutes prior to episode onset) from the individuals with T1D to identify sleep and physiological features that distinguish hypoglycaemic episodes (Level 1 and Level 2) from control nights (nights without hypoglycaemia). The non-parametric statistical testing revealed significant differences in sleep patterns and heart rate characteristics across hypoglycaemic groups. A summary of significant features by physiological category and pairwise comparison is presented in Figure 5.

**Fig 5.**
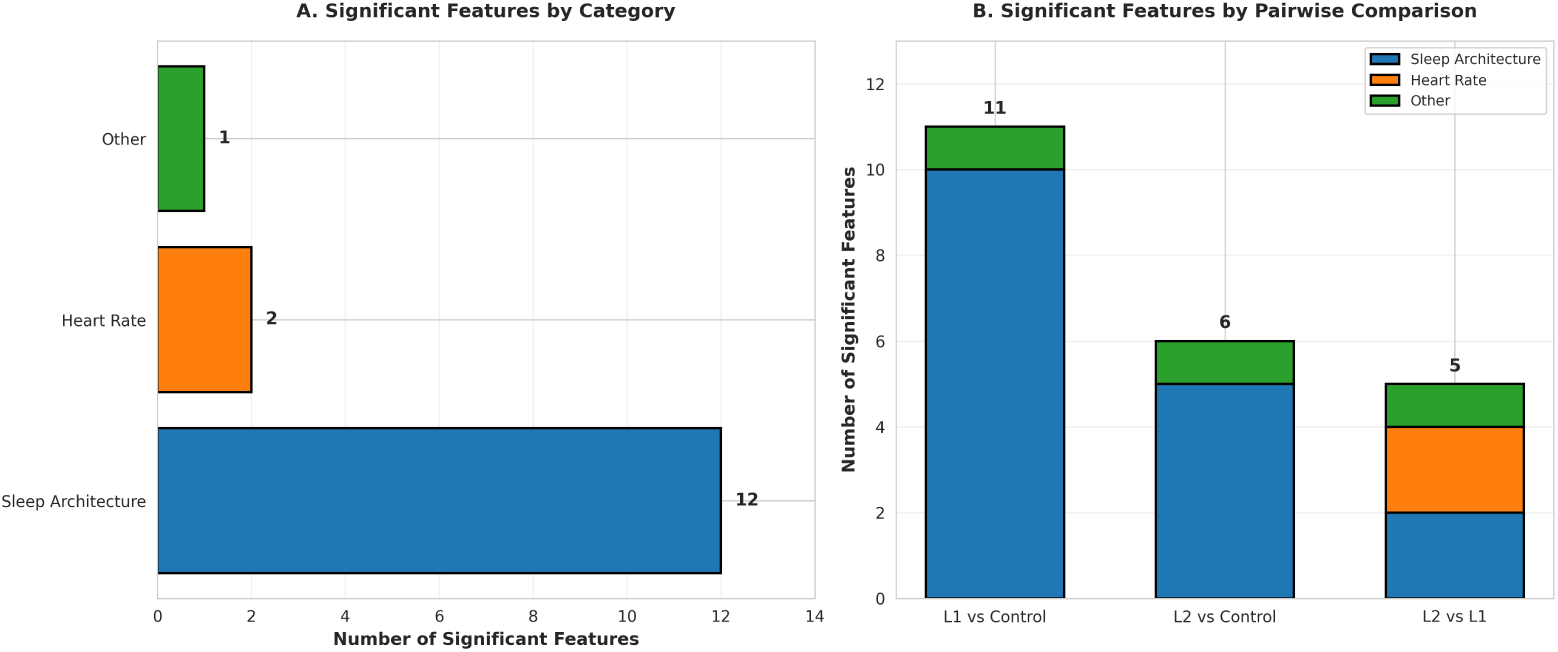
Summary of significant features by category and pairwise comparison. (A) Distribution of significant features across physiological categories, showing the predominance of sleep architecture features. (B) Breakdown of significant features by pairwise comparison (L1 vs Control, L2 vs Control, and L2 vs L1). Sleep architecture features account for the majority of significant differences, followed by heart rate and other physiological measures include step count and stress level.

Awake bouts showed the most pronounced differences between hypoglycaemic episodes and control nights (Table 4). Significantly shorter awake bouts preceded both Hypoglycaemia Level 1 and Hypoglycaemia Level 2 episodes compared to controls. Maximum awake bout duration was 79% shorter in Hypo Level 1 (2.0 min) and 74% shorter in Hypo Level 2 (3.0 min) relative to controls (11.5 min). Similarly, mean awake bout duration was reduced by 76% in Hypo Level 1 (2.0 min) and 71% in Hypo Level 2 (2.5 min) compared to controls (8.5 min).

**Table 4.** Awake bout features preceding hypoglycaemic episodes.

| Feature | Hypo L2 (n=72) | Hypo L1 (n=189) | Control (n=489) | H | q-value | Significant Comparisons |
| --- | --- | --- | --- | --- | --- | --- |
| Awake Max Bout (min) | 3.0 | 2.0 | 11.5 | 26.76 | 0.000046 | Level 1 vs Control; Level 2 vs Control |
| Awake Mean Bout (min) | 2.5 | 2.0 | 8.5 | 22.90 | 0.000209 | Level 1 vs Control; Level 2 vs Control |
| Awake Min Bout (min) | 2.0 | 2.0 | 4.0 | 9.50 | 0.039200 | Level 1 vs Control |

Abbreviations: H, Kruskal–Wallis test statistic; q-value, false discovery rate adjusted p-value.

Light sleep features exhibited distinct patterns across groups, with differences observed in both presence and bout duration (Table 5). Hypoglycaemic Level 1 episodes were preceded by an increased proportion of light sleep, accounting for 31.9% of the 60-minute pre-hypoglycaemic episode window compared to 10.8% in Hypo Level 2. In contrast, Hypoglycaemic Level 2 episodes were characterised by significantly shorter light sleep bouts. Maximum light sleep bout duration was 54% shorter in Hypo Level 2 (12.5 min) compared to controls (27.5 min), whilst mean bout duration was reduced by 49% (8.7 min vs 17.0 min). Furthermore, Hypo Level 2 episodes showed shorter light sleep bouts than Hypo Level 1, with reductions of 43% in maximum and 44% in mean bout durations.

**Table 5.** Light sleep features preceding hypoglycaemic episodes.

| Feature | Hypo L2 | Hypo L1 | Control | H | q-value | Significant Comparisons |
| --- | --- | --- | --- | --- | --- | --- |
| Light Sleep (%) | 10.8 | 31.9 | 0.0 | 22.13 | 0.000231 | Level 1 vs Control ($\Delta=31.9$) |
| Light Bout Count | 1.0 | 1.0 | 0.0 | 20.70 | 0.000377 | Level 1 vs Control ($\Delta=1.0$) |
| Light Max Bout (min) | 12.5 | 22.0 | 27.5 | 19.80 | 0.000493 | Level 2 vs Control ($\Delta=-15.0$); Level 2 vs Level 1 ($\Delta=-9.5$) |
| Light Min (min) | 5.0 | 14.0 | 0.0 | 14.51 | 0.004640 | Level 1 vs Control ($\Delta=14.0$) |
| Light Mean Bout (min) | 8.7 | 15.5 | 17.0 | 14.00 | 0.005390 | Level 2 vs Control ($\Delta=-8.3$); Level 2 vs Level 1 ($\Delta=-6.8$) |

Abbreviations: H, Kruskal–Wallis test statistic; q-value, Benjamini–Hochberg false discovery rate adjusted p-value; $\Delta$ , median difference between groups.

Heart rate was elevated prior to severe hypoglycaemic episodes (Table 6). Hypo Level 2 episodes were preceded by significantly higher heart rates compared to Hypo Level 1, with maximum heart rate 9.8% higher (95.0 bpm vs 86.5 bpm) and mean heart rate 5.7% higher (84.1 bpm vs 79.6 bpm). No significant differences were detected between Hypo Level 2 and control nights, whereas Hypo Level 1 episodes were associated with numerically lower heart rates.

**Table 6.** Heart rate features preceding hypoglycaemic episodes.

| Feature | Hypo L2 (Median) | Hypo L1 (Median) | Control (Median) | H | q-value | Significant Comparisons |
| --- | --- | --- | --- | --- | --- | --- |
| Maximum HR (bpm) | 95.0 | 86.5 | 90.0 | 11.04 | 0.0215 | Level 2 vs Level 1 (p=0.003, significant); Level 2 vs Control (p=0.0537, ns) |
| Mean HR (bpm) | 84.1 | 79.6 | 83.6 | 9.06 | 0.0423 | Level 2 vs Level 1 (p=0.008, significant); Level 2 vs Control (p=0.0632, ns) |

Abbreviations: HR, heart rate; H, Kruskal–Wallis test statistic; q-value, Benjamini–Hochberg false discovery rate adjusted p-value; bpm, beats per minute; ns, not significant.

## Discussion

This exploratory study demonstrates that NH is associated with measurable alterations in sleep architecture and autonomic physiology in free-living adults with T1D. Across both nightly and pre-hypoglycaemic episode analyses, we identified consistent patterns linking falling ISG levels with changes in sleep patterns, stage organisation, and cardiovascular activity. These results show that NH is not solely a metabolic disturbance but also a physiological and behavioural event that is measurable in wearable-derived signals long before the blood glucose level reaches its lowest point. While recent studies have demonstrated the feasibility of detecting nocturnal hypoglycaemia using wearable data [31,34], these approaches focus on classification accuracy rather than the underlying sleep and arousal mechanisms. Our findings complement this literature by providing a physiological interpretation of how sleep metrics and signals change before and during nocturnal hypoglycaemia, helping to explain why wearable-based detection is possible during sleep.

Nights with hypoglycaemic events were characterised by reduced overall wake time but increased fragmentation, alongside elevated proportions of deep and REM sleep. Specifically, while total WASO decreased, the fragmentation index, which captures brief micro-arousals and stage transitions that do not meet the wearable’s threshold for scoring a full ”awake” interval, increased substantially. This pattern indicates that these nights exhibit more frequent, shorter arousals that disrupt sleep continuity without producing extended wake periods detectable as conventional awakenings. These findings align with prior work suggesting impaired arousal and blunted counter-regulatory responses during sleep [12, 13]. However, our results extend this understanding by providing detailed, wearable-derived quantification under real-world, free-living conditions rather than controlled laboratory settings. The strong within-person effects observed across participants suggest that individuals serve as their own optimal controls, reinforcing the value of personalised modelling approaches over population-level thresholds.

One of the more unexpected patterns was the combination of shorter awake bouts and higher sleep efficiency preceding NH. In typical sleep physiology, increasing sleep efficiency reflects continuous, restorative sleep. Here, however, the 76–79% reduction in awake bout duration, compared to normo- and hyperglycaemic nights and near-complete suppression of awakening bouts likely reflect reduced arousability as blood glucose levels begin to decline. This interpretation is consistent with evidence of impaired counter-regulatory responses during sleep [13, 15] and underscores a surprising implication: increased sleep efficiency may in fact indicate elevated risk rather than improved sleep quality for people with T1D. From a clinical perspective, this pattern suggests that sleep state is a relevant factor in interpreting nocturnal blood glucose values and alarms, because individuals who appear to be “sleeping well” may, in fact, be the least likely to wake and take corrective action when hypoglycaemia develops. From hypoglycaemia detection perspective, this pattern indicates that elevated sleep efficiency, rather than representing improved sleep quality, may correspond to a state of reduced physiological and behavioural responsiveness, during which hypoglycaemic events are less likely to trigger awakening or conscious response if sleep metrics are not taken into account.

Differences between Level 1 and Level 2 hypoglycaemia provide further insight into the physiological progression of nocturnal events. Level 1 episodes were preceded by substantially more light sleep, suggesting a transitional state in which the brain maintains partial vigilance without full awakening. This aligns with the concept of cortical arousal without behavioural awakening [45]. The preservation of light sleep may reflect an intermediate stage where autonomic responses are engaged but insufficient to trigger full consciousness.

In contrast, Level 2 episodes were characterised by fragmented and shorter light-sleep bouts, coupled with a shift toward deep sleep and elevated heart rate. Critically, this pattern suggests that as blood glucose continues to decline, arousal thresholds increase rather than decrease, making it harder to wake up just when arousal would be protective. The elevated heart rate prior to Level 2 episodes likely reflects sympathetic activation driven by stronger counter-regulatory demand, although this response may still be insufficient to trigger awakening [37]. This combination of deeper sleep and increased HR without awakening may help explain why some NH events go unrecognised, even when CGM alarms are active, suggesting a possible explanation for missed alerts when sleep architecture is not considered. Both Level 1 and Level 2 events were associated with increased REM duration relative to normoglycaemic nights. Since REM sleep is characterised by autonomic instability, including rapid fluctuations in heart rate and variable sympathetic output, this increase may contribute to reduced blood glucose awareness and greater physiological vulnerability during NH. The combination of increased REM/deep-sleep, and reduced wakefulness provides a coherent physiological explanation for why NH often remains undetected.

Hyperglycaemia offered an informative contrast. While hypoglycaemia was associated with fewer awakenings but greater fragmentation, hyperglycaemia produced the most disrupted sleep overall of all three glycaemic states, with increased WASO, a higher number of awakenings, and reduced sleep quality. These opposing patterns indicate that sleep alterations during nocturnal glycaemic extremes are not uniform; hypoglycaemia leads to suppression of arousal, whereas hyperglycaemia provokes heightened disturbance. This contrast reinforces that the patterns we attribute to hypoglycaemia are not simply markers of glycaemic variability but specific features of falling blood glucose.

Pre-hypoglycaemic episode analyses revealed several early indicators of impending hypoglycaemia. Shorter awake and light-sleep bouts, increased sleep efficiency, and, during Level 2 episodes, elevated heart rate were consistently detectable in the 60-minute window prior to blood glucose decline. These changes were present even when absolute ISG was still within or near the normal range (above 4 mmol/l), highlighting their potential as early-warning physiological markers. The predominance of sleep architecture features among significant pre-hypoglycaemic episode metrics further indicates that wearable-derived sleep staging, captures clinically meaningful deviations linked to blood glucose dynamics. Taken together, these findings support the concept of a multimodal, sleep-informed algorithm that adjusts hypoglycaemia alerts according to an individual’s current sleep state, to improve sensitivity for clinically important events while minimising unnecessary sleep disruption.

Inter-individual variability in sleep timing and duration was substantial, This underscores the limitations of fixed nocturnal windows (e.g., 00:00–07:00) commonly used in glycaemic research and reinforcing the need for personalised analysis. Despite this variability, within-person changes during pre-hypoglycaemic episode periods were remarkably consistent, with effect sizes for sleep metrics consistently large within individuals despite differing baseline values. This pattern suggests that personalised, adaptive thresholds may improve hypoglycaemia detection performance. These findings have important implications for the development of multimodal detection systems that combine sleep stages, micro-awakening patterns, light-sleep fragmentation, and heart-rate dynamics. The ability of wearable devices to detect severity-specific changes before hypoglycaemia suggests that early-warning algorithms could use continuous risk levels instead of fixed thresholds. This would allow more timely and personalised interventions, helping prevent progression to severe events. For example, a gradual risk score that integrates multiple sleep features identified in this study could trigger earlier, less disruptive interventions (e.g., gentle alerts) when risk is moderate, reserving more aggressive alarms for imminent severe hypoglycaemia. Evidence from smartwatch-based detection of NH further supports the feasibility of such multimodal risk scoring approaches [28, 33]. Beyond methodological refinements, the potential of these findings is particularly relevant for low- and middle-income countries, where CGM access remains limited and NH detection is a major clinical challenge. Because consumer wearables are increasingly widespread and low cost [40], their ability to capture early physiological and behavioural signatures of hypoglycaemia could provide a scalable risk-monitoring alternative. Leveraging these signals may offer a practical step toward improving nighttime safety and reducing global disparities in T1D care.

This study has several limitations that should be taken into account. First, although the final sample size was modest, extended nightly monitoring (median 52 nights per participant) provided rich within-participant data for reliable non-parametric statistical comparisons. This depth of longitudinal data aligns with the sample sizes used in similar wearable-based T1D studies [46], supporting the robust testing of intra-individual patterns. Differences in sample characteristics, including age, diabetes duration, and hypoglycaemia awareness, may further restrict applicability to other patient groups. To address data quality concerns, this study intentionally ensured completeness and synchronisation by retaining only nights with complete CGM and sleep-stage information after applying strict preprocessing criteria (e.g., exclusion of CGM gaps exceeding 120 minutes and incomplete or unsynchronised sleep-staging outputs). However, despite the rich data per participant, validation in larger and more diverse cohorts remains necessary to establish the generalisability of these findings. A second limitation relates to the sleep-staging accuracy of consumer wearable devices. As the study relied on wearable-derived staging rather than polysomnography, the state of the art in sleep studies, the Garmin smartwatch algorithms may introduce misclassification of sleep stages, notably in distinguishing between light and deep sleep, which could influence the precision of specific stage-level metrics [36, 43, 44]. To mitigate these device-related uncertainties, we implemented several quality-focused preprocessing procedures, including retaining only complete nights, carefully aligning multimodal signals, and inspecting multiple conditions (e.g., timestamp synchronisation, sleep onset, and episode-window boundaries). Additionally, we computed a deeper level of derived sleep features, including stage fragmentation indices, stability indices, and detailed transition counts analysed by hypoglycaemia severity. These aggregate metrics may be more robust to individual epoch misclassification than raw stage durations alone. However, we acknowledge that any staging inaccuracies may still have influenced the resolution of these metrics. Future validation studies comparing wearable-derived staging against concurrent polysomnography in individuals with T1D would help quantify the magnitude of this potential bias and strengthen confidence in these findings.

Third, the analysis focused on a single 60-minute pre-hypoglycaemic episode window, which may not capture longer-term or cumulative changes that precede hypoglycaemia. To partially address these concerns, we included heart rate and stress metrics derived from wearable sensors, providing some insight into physiological state during the pre-hypoglycaemic episode window. However, longer observation windows, potentially spanning multiple nights or wake periods, may reveal additional predictive patterns. What is more, unmeasured confounders such as alcohol consumption, medication timing, stress, physical activity, and meal composition could have influenced both sleep and blood glucose dynamics. Other physiological parameters, such as heart rate variability, skin temperature, and respiratory rate, were not analysed and may offer further insight [47,48]. The absence of direct mechanistic measures, including counter-regulatory hormones, autonomic activity, and cortical arousal, prevents definitive conclusions about underlying physiological pathways. Future research incorporating expanded physiological sensing and longer observation windows would help clarify the temporal dynamics and mechanistic pathways linking sleep disruption to NH.

Finally, building on these findings, future research should validate the identified sleep features in larger and more diverse cohorts, incorporating multiple wearable devices to cross-validate sleep staging against polysomnography. Algorithmic work is needed to translate these features into real-time detection systems, particularly those using adaptive, within-person baselines. Evaluations in low-resource environments will be essential to assess feasibility, acceptability, and impact on nocturnal safety. Together, these steps will be critical for realising the potential of sleep-informed digital technologies to improve NH detection and prevention.

## Materials and methods

### Ethics Statement

Ethical approval was granted by the University of Manchester Research Ethics Committee (Ref: 2023-15687-29584) and conducted in accordance with the Declaration of Helsinki and Good Clinical Practice guidelines. The study used a longitudinal observational approach, collecting data between October 1, 2023, and September 3, 2024. All participants provided written informed consent and were given at least 24 hours to consider participation before enrolment.

### Study Design and Data Collection

This study employed a retrospective analysis of CGM data paired with Garmin smart-watch sleep-tracking information from a publicly available dataset reported in Nature Scientific Data [49]. Researchers enrolled participants via multiple channels: the IAM Lab’s social media platforms, Type 1 diabetes-focused online communities, email campaigns, Twitter posts, various social media announcements, and printed flyers distributed across University of Manchester buildings and surrounding areas including local sports facilities. Data collection and devices timestamps were standardised across participants, with CGM and smartwatch-derived physiological signals recorded throughout the day. Nocturnal monitoring was prioritised to enable synchronised acquisition of blood glucose dynamics and sleep parameters. Participants were instructed to maintain their usual lifestyle habits and refrain from altering their daily routines to preserve ecological validity. Raw sensor-derived blood glucose readings, referred to as interstitial blood glucose (ISG) values, and sleep data were imported for data manipulation and analysis. For each participant, the dataset contained blood glucose readings with corresponding timestamps, alongside rich sleep data derived from the smartwatch. Sleep information included physiological markers (HR, stress level, and step count), as well as sleep features such as sleep episode start and end times, metadata on sleep stages, and summary measures reporting total time spent in each stage (Deep, REM, Light, Awake) and overall total sleep time in seconds. The structure and content of the dataset are illustrated in Figure 6.

**Fig 6.**
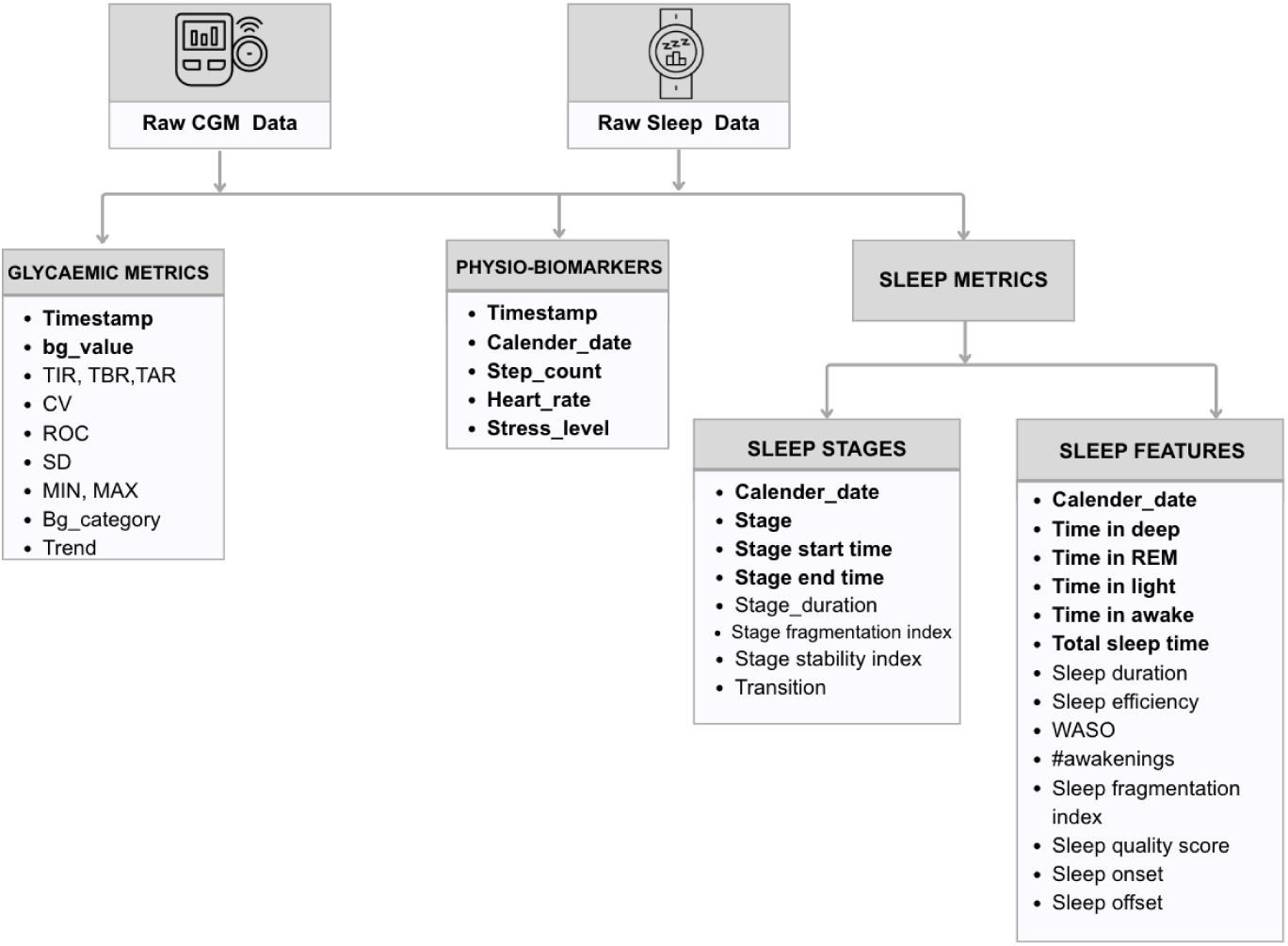
Structure of the dataset combining continuous glucose monitoring (CGM) metrics with smartwatch-derived sleep data, including physiological biomarkers, stage-level information, and summary sleep features.

### Participants

Seventeen adults with T1D were recruited and monitored for 12 weeks. During the initial interview, participants consented to access their blood glucose sensor and insulin device platforms. Additional consent was obtained at the end of the study period for access to smartwatch data. Combined CGM and sleep-tracking data were available for 14 participants. Participants were excluded if their data showed (1) repeated CGM gaps *>*120 minutes during a night, (2) missing or incomplete sleep-stage outputs, or (3) unsynchronised timestamps preventing accurate CGM–sleep alignment. After applying these criteria, 11 participants (573 nights) were included in the final analysis, as summarised in Table 7. All participants consented to the publication of their anonymised data.

**Table 7.** Participant demographics.

| Participant ID | Sex | Age range (years) | Sensor | TIR (%) |
| --- | --- | --- | --- | --- |
| UoM2302 | F | 25–29 | Flash | 87.81 |
| UoM2303 | F | 25–29 | CGM | 92.92 |
| UoM2304 | F | 25–29 | CGM | 71.17 |
| UoM2306 | F | 50–54 | Flash | 82.83 |
| UoM2307 | F | 60–64 | CGM | 67.80 |
| UoM2308 | M | 55–59 | CGM | 84.41 |
| UoM2309 | F | 55–59 | CGM | 54.29 |
| UoM2313 | M | 35–39 | Flash | 52.97 |
| UoM2320 | F | 45–49 | CGM | 93.88 |
| UoM2401 | M | 45–49 | Flash | 70.16 |
| UoM2404 | F | 35–39 | Flash | 71.37 |

### Data processing

Data processing involved several steps to ensure consistency across participants and accurate alignment of ISG and sleep data. Both CGM and smartwatch data were resampled to 15-minute intervals to achieve consistent temporal alignment across all participants with T1D. Night records were excluded if they contained gaps in ISG measurements exceeding 120 minutes or if sleep information was missing. When clock shifts due to time zone differences (e.g., daylight saving time transitions or travel) were detected, timestamps were adjusted accordingly.

To align with the literature on NH, nighttime was defined as 00:00–07:00 [50, 51]. We used this window to confirm that participants adhered to conventional nocturnal sleep schedules (i.e., were not shift workers). The sleep periods used in this study were derived from smartwatch-recorded sleep onset and offset times and could overlap with, fall within, or extend beyond this fixed nighttime interval. Sleep and ISG data were integrated using a date-based matching approach, whereby sleep onset times and ISG timestamps were used to align glycaemic events with corresponding sleep periods. Data merging was performed using participant identifiers and date as matching keys, and only nights with complete sleep and ISG data were retained for analysis. An overview of the methodological workflow, including data collection, preprocessing, feature derivation, and analytical steps, is presented in Figure 7.

**Fig 7.**
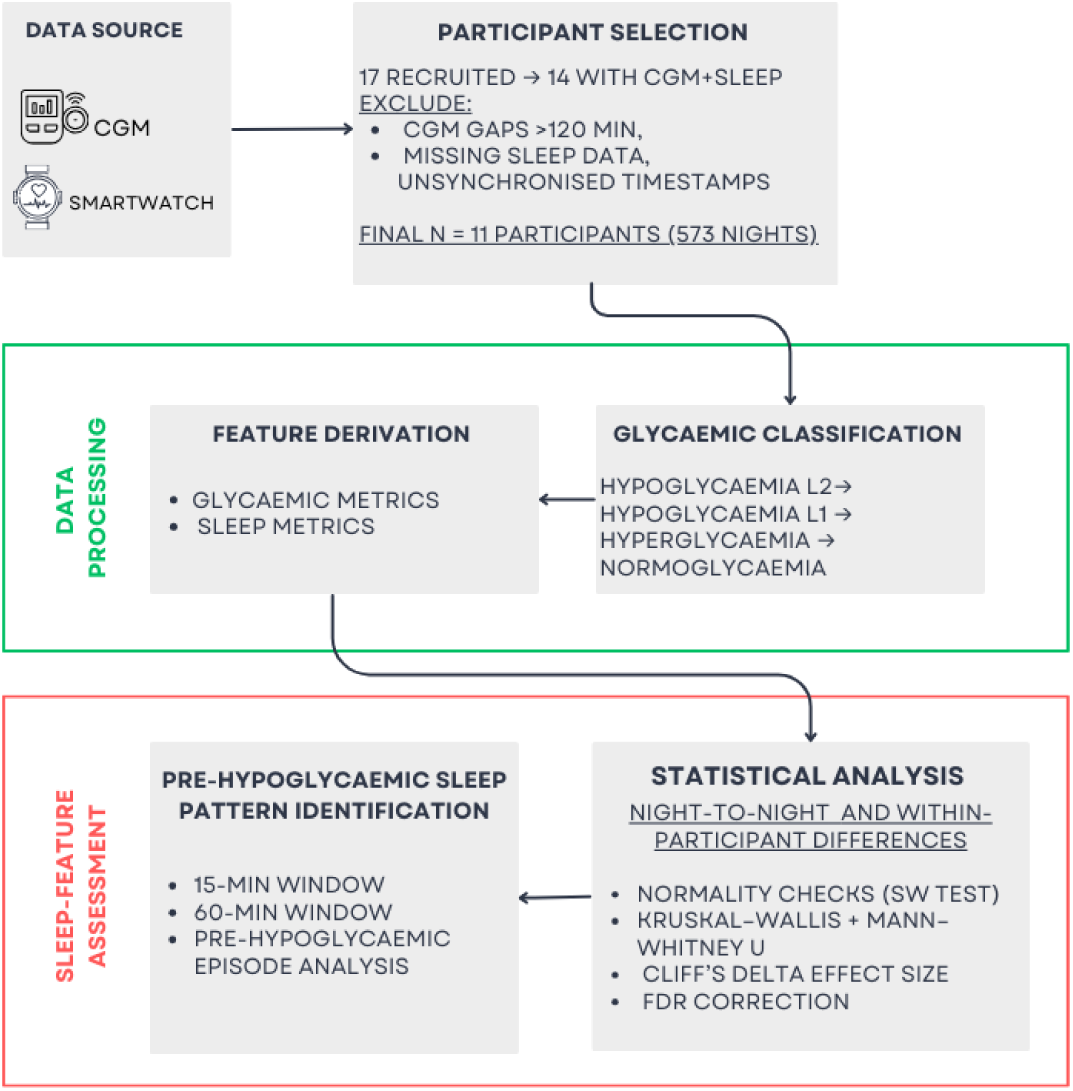
Major methodological steps of the study, summarising data collection, preprocessing, feature derivation, glycaemic classification, and statistical analyses.

Glycaemic events were classified into four hierarchical categories based on consensus clinical thresholds: Normoglycaemia (4.0–10.0 mmol/L), Hyperglycaemia (*>*10.0 mmol/L), Hypoglycaemia Level 1 (*≥*3.1 and *<*4.0 mmol/L), and Hypoglycaemia Level 2 (*<*3.1 mmol/L). For nights with multiple glycaemic events, a precedence-based classification system was applied to assign a single clinically meaningful category per participant-night, prioritising the most severe event in the following order: Hypoglycaemia Level 2 *→* Hypoglycaemia Level 1 *→* Hyperglycaemia *→* Normoglycaemia. This hierarchical approach ensured that each participant-night received exactly one glycaemic classification reflecting the most clinically significant event of that night.

### Feature Derivation

#### Glycaemic metrics

Glycaemic metrics (Figure 6) were derived from ISG readings after resampling and temporal alignment to sleep period timestamps. For each participant-night, summary statistics included the mean and standard deviation of ISG values during the sleep period. Blood glucose variability was quantified using two complementary measures: the coefficient of variation (CV), calculated as (standard deviation/mean) *×* 100%, and the rate of change (ROC), computed as the first derivative between consecutive ISG measurements (mmol/L per 15 minutes) to capture blood glucose fluctuation trends.

Clinical glycaemic ranges were assessed by calculating both the absolute duration (minutes) and relative proportion (%) of time spent in three key states: time in range (TIR: 3.9–10.0 mmol/L), time below range (TBR: *<*3.9 mmol/L), and time above range (TAR: *>*10.0 mmol/L). These metrics were calculated relative to the total duration of the sleep period. Complete definitions of all glycaemic features are provided in Supplementary Table SB.

#### Sleep metrics

Sleep features were derived at two levels of granularity: stage-level metrics characterising within-stage dynamics, and nightly aggregated metrics summarising overall sleep architecture. All metrics were calculated per participant-night.

*Nightly aggregated metrics* included standard sleep parameters: total sleep time (TST), sleep efficiency (TST as the total time in bed, including arousal time), wake after sleep onset (WASO), and the percentage and relative durations of each sleep stage (Light, Deep, REM). Two composite indices were derived to quantify overall sleep quality:

- The Sleep Fragmentation Index (SFI) assessed sleep continuity by calculating the total number of nocturnal awakenings and sleep stage transitions, normalised by TST in hours:

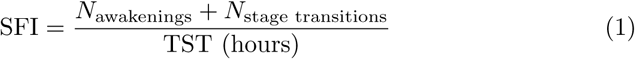

where *N*_stage transitions_ includes all transitions between Light, Deep, and REM stages. This index was adapted from an actigraphy-based definition described previously by Knutson et al. (2011) [10]. In the wearable-derived sleep architecture, fragmentation indices include all stage transitions and micro-arousals, including those too brief (less than 5 minutes) to be classified as full awake epochs by the device’s staging algorithm [43, 52, 53].

- The Sleep Quality Score (SQS) was computed as a custom composite metric, scaled from 0 to 100, to provide an overall assessment of sleep quality using the smartwatch-derived sleep measures available in this study. This approach was informed by prior composite scoring strategies used in wearable-based sleep research and existing recommendations on sleep assessment, for which a standardised composite metric is not yet available [53–55, 55]. The SQS incorporated the following normalised and weighted components of sleep architecture and continuity: sleep efficiency (25%), sleep duration (20%), WASO (6%), number of awakenings (4%), REM sleep percentage (10%), deep sleep percentage (10%), and SFI (25%). Absolute values of these components may vary depending on the smartwatch algorithm used for sleep staging and event detection. Each component was first normalised to a 0-100 scale before applying the respective weights, with higher scores indicating better overall sleep quality.

Following established approaches in prior sleep research, *Stage-level metrics* were derived to characterise the micro-architecture and continuity of individual sleep stages through four complementary measures:

1. **Stage Fragmentation Index (GFI):** The ratio of stage transitions to total sleep intervals within each stage, quantifying the degree of architectural disruption (expressed as transitions per interval).
2. **Stage Stability Index (SSI):** The mean duration (minutes) of continuous, uninterrupted bouts within each stage, reflecting stage persistence and resistance to fragmentation.
3. **Stage Bout Duration:** The median continuous time (minutes) spent in each stage before transitioning to another stage, providing a robust measure of typical stage persistence. Calculated separately for Light, Deep, and REM stages.
4. **Stage Transition Analysis:** A comprehensive characterisation of movement between sleep stages, including transition frequencies (counts) and cumulative durations (minutes) for all stage-to-stage transitions (e.g., Light*→*Deep, Deep*→*REM, REM*→*Light).

Complete definitions and computational details for all sleep features are provided in Supplementary Table SA.

### Statistical Analysis

The derived glycaemic and sleep features were subjected to comprehensive statistical analysis to identify metrics that exhibited significant differences across glycaemic event classifications and could serve as potential indicators for detecting NH.

Descriptive statistics (mean and standard deviation) were computed for all features within each night classification based on glycaemic events, with variability measures (e.g., standard deviation of sleep onset times) used to quantify sleep timing and regularity. Shapiro-Wilk tests were performed to assess distributional assumptions and indicated significant deviations from normality across all key variables (*p <* 0.001), including ISG values, heart rate, step count, stress level, and sleep-related features. Given these violations of normality assumptions, non-parametric statistical methods were applied throughout all subsequent analyses. This approach ensured that statistical inference was robust to outliers, which are common in longitudinal wearable-derived physiological data.

Group comparisons between glycaemic event classifications were conducted using the Kruskal-Wallis test [56], as comparisons involved more than two independent glycaemic classifications, and the variables violated normality assumptions. Accordingly, the Kruskal-Wallis test, a non-parametric alternative to one-way ANOVA, was used to assess overall differences across the four night classifications for each feature. For features demonstrating significant overall group differences (*p <* 0.05), post-hoc pairwise comparisons were performed using the Mann–Whitney U test to identify specific between-group differences. Effect sizes were quantified using Cliff’s delta [57], which provides a robust measure of the magnitude and direction of differences between distributions, with values interpreted as negligible (*|δ| <* 0.147), small (0.147 *≤ |δ| <* 0.33), medium (0.33 *≤ |δ| <* 0.474), or large (*|δ| ≥* 0.474). All significance tests were two-sided. Statistical analyses were performed in Python.

To account for individual variability in sleep and ISG regulation while identifying features with consistent discriminatory capacity for NH, the analytical framework incorporated two complementary approaches:

1. **Night-to-Night Analysis:** Examined inter-night variability across all participants by pooling data from all participant-nights. This population-level analysis identified generalisable patterns and assessed the consistency of sleep–glucose relationships across the cohort, revealing features that consistently differentiate hypoglycaemic nights from other glycaemic states at the group level.
2. **Within-Participant Analysis:** Focused on intra-individual differences by conducting separate statistical analyses for each participant independently. This approach identified features that showed consistent directional changes associated with hypoglycaemia within individuals, accounting for person-specific baseline differences in sleep architecture and ISG dynamics. Only participants with sufficient numbers of nights across glycaemic classifications were included in within-participant analyses.

Features demonstrating significant discriminatory capacity in both analytical approaches were considered robust candidate indicators for NH detection, as they exhibited both population-level generalisability and within-individual consistency.

### Pre-Hypoglycaemic Sleep Pattern Identification

Building upon the identification of features that distinguish hypoglycaemic nights from other glycaemic states, the analysis was further refined to characterise the temporal dynamics of physiological changes immediately preceding hypoglycaemic episodes. This approach aimed to identify early warning signatures within the pre-hypoglycaemic episode period that could inform real-time detection systems.

#### Pre-hypoglycaemic episode window definition and feature extraction

To capture the dynamic progression toward hypoglycaemia while accounting for both gradual and acute physiological changes, features were extracted from two temporally distinct pre-hypoglycaemic episode windows:

- **Long window:** 60 minutes immediately preceding hypoglycaemic episode onset
- **Short window:** 15 minutes immediately preceding hypoglycaemic episode onset

These windows were chosen based on physiological and algorithmic considerations. Autonomic changes (elevated heart rate, altered arousal) begin 30–60 minutes before hypoglycaemic events [58], so the 60 minute window captures gradual drift while the 15 minute window captures rapid pre-event changes. Wearables sample heart rate every 30–120 seconds and sleep stages every 60–120 seconds [59], making these windows sufficient for robust analysis despite missing data. Both windows were temporally anchored to the episode onset time and bounded by sleep onset to ensure all analysed data occurred during the sleep period (Figure 8). When an episode occurred within 60 minutes of sleep onset, the long window was truncated to begin at sleep onset; the same truncation rule applied to the short window if the episode occurred within 15 minutes of sleep onset. Episodes occurring within 15 minutes of sleep onset were excluded from pre-hypoglycaemic episode analysis due to insufficient pre-event data. If multiple episodes occurred within the same night, each episode was treated as an independent pre-hypoglycaemic episode event only if its pre-hypoglycaemic episode windows did not overlap with the preceding episode’s windows. Overlapping events were merged and treated as a single extended hypoglycaemic event, preventing artificial inflation of sample counts and avoiding correlated observations. All sleep architecture metrics, physiological parameters, and blood glucose metrics were computed independently for each window.

**Fig 8.**
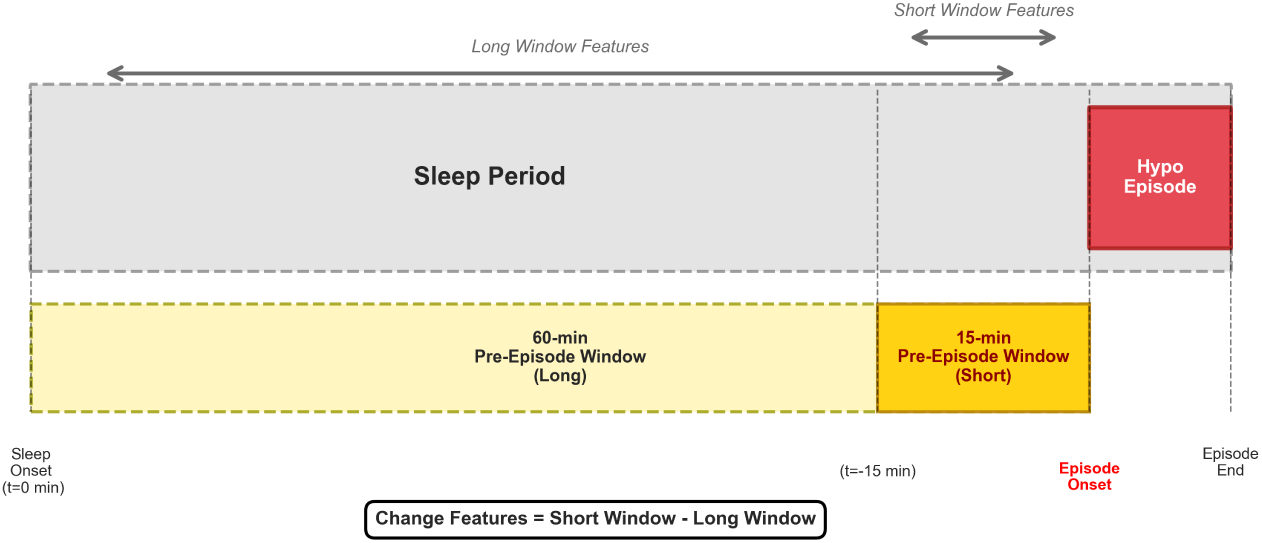
Pre-hypoglycaemic episode window analysis timeline. Illustration of the 60-minute (long) and 15-minute (short) pre-hypoglycaemic episode windows used for feature extraction before hypoglycaemic episodes. Both windows were temporally aligned relative to episode onset and bounded by sleep onset to ensure that all analysed data occurred during the sleep period. The non-overlapping design enabled assessment of both sustained pre-hypoglycaemic episode patterns and acute changes immediately before episode onset.

To quantify the acceleration or deceleration of physiological and sleep-related changes as hypoglycaemia approached, temporal change features were computed by comparing measurements between the two pre-hypoglycaemic episode windows:

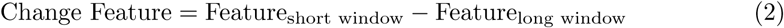

These change features were calculated for all sleep architecture, physiological, and blood glucose metrics, representing the directional shift between the 60-minute and 15-minute pre-hypoglycaemic episode windows. Positive change values indicated an increase in the feature as the episode approached, while negative values indicated a decrease. This temporal derivative approach enabled detection of accelerating trends (e.g., rapidly declining ISG, increasing heart rate) that may signal imminent hypoglycaemia more sensitively than absolute values alone.

#### Pre-hypoglycaemic episode analysis

To identify features that distinguish level 2 hypoglycaemia from level 1 hypoglycaemia during the pre-hypoglycaemic episode period, comparative analyses were performed using features derived from the 60-minute pre-hypoglycaemic episode window. The longer window was selected to capture sustained physiological differences that may precede episodes of varying severity, rather than acute changes that could be artifacts of the episode itself. In this analysis, nights without hypoglycaemia during the sleep period were used as control nights, and comparisons were restricted to hypoglycaemic episodes (L1, L2) to establish baseline sleep and physiological patterns.

Group comparisons were conducted using the Kruskal–Wallis H-test to assess overall differences between Level 1 and Level 2 episodes. For features demonstrating significant overall differences, pairwise comparisons were performed using Dunn’s post-hoc test to control the family-wise error rate [60]. To ensure statistical reliability, only features with data available for at least five episodes in both the Level 1 and Level 2 groups were included in the analysis, as smaller sample sizes may yield unstable estimates.

Effect sizes were quantified as the median difference between Level 2 and Level 1 episodes, providing a clinically interpretable measure of the magnitude of between-group differences. To control for multiple testing across the numerous features examined, *p*-values were adjusted using the Benjamini–Hochberg False Discovery Rate (FDR) procedure [61], which maintains an appropriate balance between Type I error control and statistical power. Statistical significance was defined as an FDR-adjusted *q* -value of less than 0.05, indicating that fewer than 5% of features identified as significant were expected to be false discoveries.

Features demonstrating significant differences between hypoglycaemia levels in the pre-hypoglycaemic episode window were considered candidate biomarkers for severity stratification, potentially enabling more targeted intervention strategies based on anticipated episode severity.

## Conclusion

This study provides evidence that NH is associated with distinct and measurable alterations in sleep architecture and autonomic physiology in free-living adults with T1D. By combining CGM with multimodal wearable data, we show that both the onset and level of NH are preceded by characteristic changes in sleep continuity, light and deep sleep organisation, and heart rate dynamics. These findings demonstrate that hypoglycaemia is not only a metabolic event but also a detectable physiological state that emerges well before blood glucose reaches clinically low thresholds. The notable increase in deep sleep during Level 2 episodes suggests reduced behavioural arousal at the time when awakening would be most protective, which may limit the effectiveness of conventional CGM alarms during severe NH.

Integrating wearable derived sleep-stage information with blood glucose signals may therefore enhance the reliability of nocturnal hypoglycaemia detection, particularly by reducing missed alerts when individuals enter deeper sleep states. Such sleep-informed approaches may enable more personalised and adaptive early-warning systems capable of distinguishing between nocturnal events, thereby supporting safer nighttime management for people with T1D.

Further validation in larger and more diverse populations, alongside the integration of additional physiological features and real-time algorithmic development, will be essential for translating these findings into actionable early warning tools. Nonetheless, this work establishes introductory evidence that sleep and physiological signals captured by affordable wearables can provide meaningful insight into NH and its progression, supporting a new avenue for precision digital diabetes care.

## Data Availability

The dataset is available in a Zenodo repository ‘T1D-UOM – A Longitudinal Multimodal Dataset of Type 1 Diabetes’ at https://doi.org/10.5281/zenodo.15806142.

## Acknowledgments

This research was supported by Taibah University (TaibahU). The authors extend their gratitude to all participants who took part in this study. Their willingness to share personal health, sleep, and blood glucose monitoring data over an extended period was essential to this research.

## Supporting information

**Table SA.** Smartwatch-derived sleep variables: definitions.

| Feature name | Unit | Definition |
| --- | --- | --- |
| participant_id | – | Participant identifier |
| sleep_start_time | datetime | Start of sleep as recorded by the smartwatch |
| sleep_end_time | datetime | End of sleep as recorded by the smartwatch |
| calendar_date | date | Anchor date for the night |
| Deep (min) | minutes | Total duration of Deep sleep intervals |
| Light (min) | minutes | Total duration of Light sleep intervals |
| REM (min) | minutes | Total duration of REM sleep intervals |
| Awake (min) | minutes | Total duration of Awake intervals |
| Time in Deep (%) | % | Proportion of sleep spent in Deep stage |
| Time in Light (%) | % | Proportion of sleep spent in Light stage |
| Time in REM (%) | % | Proportion of sleep spent in REM stage |
| Wake after sleep onset (min) | minutes | Total Awake time after the first non-Awake stage |
| Number of awakenings (N_awakening) | count | Count of awakenings during sleep |
| Total sleep time (TST) | minutes | Total sleep including Awake intervals |
| Sleep duration | minutes | Total duration of sleep excluding Awake intervals |
| Sleep_efficiency | % | Sleep duration / Total sleep time |
| sleep_onset | datetime | First sleep stage excluding Awake |
| awake_offset | datetime | Last sleep stage before prolonged wake |
| Intervals counted (N_interval) | count | Number of recorded stage intervals |
| deep_sleep_s | seconds | Total Deep sleep in seconds (device output) |
| light_sleep_s | seconds | Total Light sleep in seconds (device output) |
| rem_sleep_s | seconds | Total REM sleep in seconds (device output) |
| awake_sleep_s | seconds | Total Awake time in seconds (device output) |
| Sleep_duration [summary_s] | minutes | Summary sleep duration excluding Awake |
| Total sleep time (TSTs) | minutes | Summary total sleep including Awake |
| Time in Deep [summary_s] | % | Proportion of summary Deep sleep |
| Time in Light [summary_s] | % | Proportion of summary Light sleep |
| Time in REM [summary_s] | % | Proportion of summary REM sleep |
| Sleep Quality Score (SQS) | 0–100 | Composite metric of overall sleep quality |
| Sleep Fragmentation Index (SFI) | events/h | Fragmentation based on awakenings and stage transitions |
| Stage Fragmentation Index (GFI) | transitions/interval | Stage transitions per interval within a stage |
| Stage Stability Index (SSI) | minutes | Mean uninterrupted duration of each sleep stage |
| Stage Bout Duration | minutes | Median continuous time spent within a stage (Light/Deep/REM) |
| Stage Transition Analysis | counts/minutes | Frequency and duration of transitions between stages |

**Table SB.**
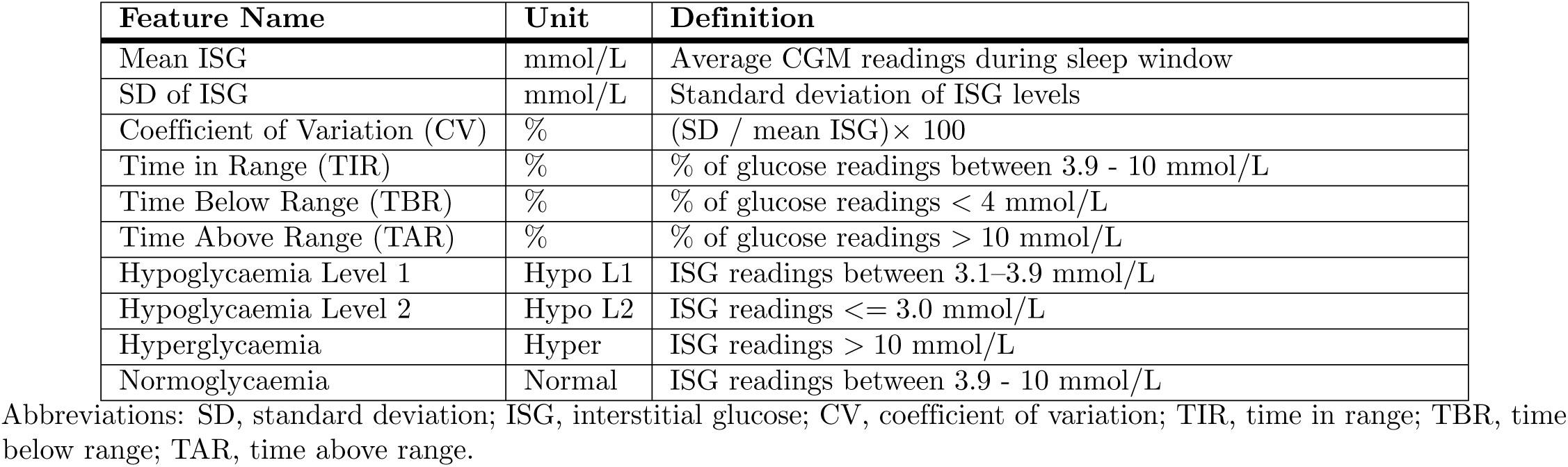
CGM-derived glycaemic features calculated during the sleep window.

| Feature Name | Unit | Definition |
| --- | --- | --- |
| Mean ISG | mmol/L | Average CGM readings during sleep window |
| SD of ISG | mmol/L | Standard deviation of ISG levels |
| Coefficient of Variation (CV) | % | (SD / mean ISG) × 100 |
| Time in Range (TIR) | % | % of glucose readings between 3.9 - 10 mmol/L |
| Time Below Range (TBR) | % | % of glucose readings < 4 mmol/L |
| Time Above Range (TAR) | % | % of glucose readings > 10 mmol/L |
| Hypoglycaemia Level 1 | Hypo L1 | ISG readings between 3.1–3.9 mmol/L |
| Hypoglycaemia Level 2 | Hypo L2 | ISG readings ≤ 3.0 mmol/L |
| Hyperglycaemia | Hyper | ISG readings > 10 mmol/L |
| Normoglycaemia | Normal | ISG readings between 3.9 - 10 mmol/L |

Abbreviations: SD, standard deviation; ISG, interstitial glucose; CV, coefficient of variation; TIR, time in range; TBR, time below range; TAR, time above range.

**Table SC.**
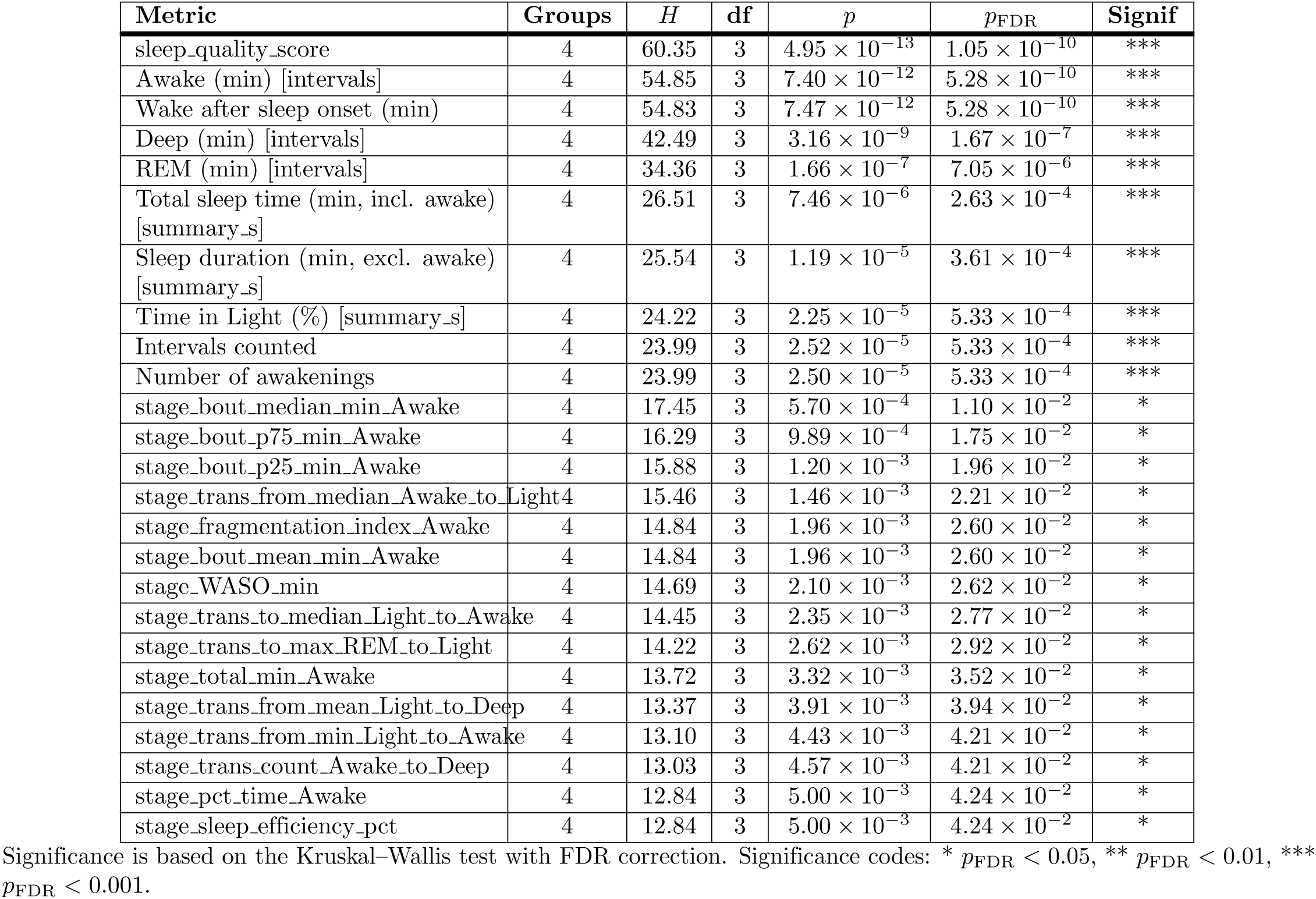
Kruskal–Wallis test results for sleep metrics with significant group differences.

**Table SD.** Pairwise comparisons of sleep metrics across nocturnal glycaemic states.

| Metric | Group A | Median A | Mean A | Group B | Median B | Mean B | $p_{\text{FDR}}$ |
| --- | --- | --- | --- | --- | --- | --- | --- |
| Awake (min) | Hypo Level 2 | 42 | 57.88 | Hyper | 73 | 96.53 | 1.04e-08 |
| Awake (min) | Hypo Level 1 | 49 | 66.56 | Hyper | 73 | 96.53 | 3.31e-08 |
| Awake (min) | Hypo Level 2 | 42 | 57.88 | Normal | 64 | 75.34 | 0.000485 |
| Awake (min) | Hypo Level 1 | 49 | 66.56 | Normal | 64 | 75.34 | 0.0148 |
| Awake (min) | Hyper | 73 | 96.53 | Normal | 64 | 75.34 | 0.043 |
| Sleep duration (min) | Hypo Level 2 | 477 | 471.29 | Hyper | 458 | 450.20 | 0.0199 |
| Sleep duration (min) | Hypo Level 1 | 467 | 458.47 | Hyper | 458 | 450.20 | 0.144 |
| Sleep duration (min) | Hypo Level 2 | 477 | 471.29 | Normal | 454.5 | 455.03 | 0.0592 |
| Sleep duration (min) | Hypo Level 1 | 467 | 458.47 | Normal | 454.5 | 455.03 | 0.777 |
| Sleep duration (min) | Hyper | 458 | 450.20 | Normal | 454.5 | 455.03 | 0.279 |
| Sleep efficiency (%) | Hypo Level 2 | 84.07 | 83.53 | Hyper | 79.74 | 79.08 | 0.000163 |
| Sleep efficiency (%) | Hypo Level 1 | 83.75 | 82.67 | Hyper | 79.74 | 79.08 | 2.76e-08 |
| Sleep efficiency (%) | Hypo Level 2 | 84.07 | 83.53 | Normal | 81.76 | 81.71 | 0.00312 |
| Sleep efficiency (%) | Hypo Level 1 | 83.75 | 82.67 | Normal | 81.76 | 81.71 | 0.000166 |
| Sleep efficiency (%) | Hyper | 79.74 | 79.08 | Normal | 81.76 | 81.71 | 0.00281 |
| WASO (min) | Hypo Level 2 | 84 | 88.38 | Hyper | 117 | 127.76 | 0.000141 |
| WASO (min) | Hypo Level 1 | 89 | 97.09 | Hyper | 117 | 127.76 | 2.60e-08 |
| WASO (min) | Hypo Level 2 | 84 | 88.38 | Normal | 101 | 103.23 | 0.00314 |
| WASO (min) | Hypo Level 1 | 89 | 97.09 | Normal | 101 | 103.23 | 0.000151 |
| WASO (min) | Hyper | 117 | 127.76 | Normal | 101 | 103.23 | 0.00267 |
| REM duration (min) | Hypo Level 2 | 87 | 86.46 | Hyper | 62 | 68.42 | 4.68e-06 |
| REM duration (min) | Hypo Level 1 | 78 | 80.26 | Hyper | 62 | 68.42 | 2.13e-05 |
| REM duration (min) | Hypo Level 2 | 87 | 86.46 | Normal | 71 | 73.62 | 6.15e-05 |
| REM duration (min) | Hypo Level 1 | 78 | 80.26 | Normal | 71 | 73.62 | 0.000655 |
| REM duration (min) | Hyper | 62 | 68.42 | Normal | 71 | 73.62 | 0.0019 |
| REM proportion (%) | Hypo Level 2 | 18.40 | 18.47 | Hyper | 14.11 | 14.95 | 1.84e-05 |
| REM proportion (%) | Hypo Level 1 | 16.80 | 17.50 | Hyper | 14.11 | 14.95 | 5.65e-05 |
| REM proportion (%) | Hypo Level 2 | 18.40 | 18.47 | Normal | 15.20 | 15.90 | 6.32e-05 |
| REM proportion (%) | Hypo Level 1 | 16.80 | 17.50 | Normal | 15.20 | 15.90 | 0.00421 |
| REM proportion (%) | Hyper | 14.11 | 14.95 | Normal | 15.20 | 15.90 | 0.0234 |
| Number of awakenings | Hypo Level 2 | 10 | 11.13 | Hyper | 13 | 14.90 | 0.00114 |
| Number of awakenings | Hypo Level 1 | 11 | 11.94 | Hyper | 13 | 14.90 | 1.91e-05 |
| Number of awakenings | Hypo Level 2 | 10 | 11.13 | Normal | 12 | 13.03 | 0.00391 |
| Number of awakenings | Hypo Level 1 | 11 | 11.94 | Normal | 12 | 13.03 | 0.00112 |
| Number of awakenings | Hyper | 13 | 14.90 | Normal | 12 | 13.03 | 0.0162 |
| Average awakening duration (min) | Hypo Level 2 | 7 | 8.37 | Hyper | 8 | 9.00 | 0.0931 |
| Average awakening duration (min) | Hypo Level 1 | 8 | 9.05 | Hyper | 8 | 9.00 | 0.557 |
| Average awakening duration (min) | Hypo Level 2 | 7 | 8.37 | Normal | 7 | 8.39 | 0.781 |
| Average awakening duration (min) | Hypo Level 1 | 8 | 9.05 | Normal | 7 | 8.39 | 0.163 |
| Average awakening duration (min) | Hyper | 8 | 9.00 | Normal | 7 | 8.39 | 0.134 |

Pairwise comparisons are based on Dunn's post-hoc test with false discovery rate (FDR) correction. Effect size classification (negligible / small / medium) is based on Cliff's delta.

